# PALM: Patient-centered Treatment Ranking via Large-scale Multivariate Network Meta-analysis

**DOI:** 10.1101/2020.11.18.20234252

**Authors:** Rui Duan, Jiayi Tong, Lifeng Lin, Lisa D Levine, Mary D Sammel, Joel Stoddard, Tianjing Li, Christopher H Schmid, Haitao Chu, Yong Chen

## Abstract

The growing number of available treatment options have led to urgent needs for reliable answers when choosing the best course of treatment for a patient. As it is often infeasible to compare a large number of treatments in a single randomized controlled trial, multivariate network meta-analyses (NMAs) are used to synthesize evidence from existing trials of a subset of the available treatments, where outcomes related to both efficacy and safety are considered simultaneously. However, these large-scale multiple-outcome NMAs have created challenges to existing methods due to the increasingly complexity of the unknown correlation structures between different outcomes and treatment comparisons. In this paper, we proposed a new framework for PAtient-centered treatment ranking via Large-scale Multivariate network meta-analysis, termed as PALM, which includes a parsimonious modeling approach, a fast algorithm for parameter estimation and inference, a novel visualization tool for comparing treatments with multivariate outcomes termed as the star plot, as well as personalized treatment ranking procedures taking into account the individual’s considerations on multiple outcomes. In application to an NMA that compares 14 treatment options for labor induction over five modalities, we provided a comprehensive illustration of the proposed framework and demonstrated its computational efficiency and practicality. Our analysis leads to new insights on comparing these 14 treatment options based on joint inference of multiple outcomes that cannot be obtained from univariate NMAs, and novel visualizations of evidence to support patient-centered clinical decision making.

## 1 Introduction

In the areas of evidenced-based medicine and clinical decision making, clinicians are now facing challenges caused by the growing number of available treatment options for a specific condition. For example, there are over 20 commonly prescribed treatments for major depressive disorder, and a comparison between all treatments in a single randomized controlled trial (RCT) is infeasible, making it difficult for clinicians to draw conclusions regarding the relative effectiveness of all treatments (Cipriani et al. 2018). In addition to the increasing number of available treatments, clinicians, patients and stakeholders often need to consider multiple aspects when choosing a treatment. Specifically, in addition to a primary outcome that often reflects treatment efficacy, there are secondary outcomes that are also important in clinical decision making, such as adverse effects, quality of life outcomes, long-term risks, among others (Davey et al. 2011). In sum, the proliferation of treatment options for a condition has increased the complexity of clinical decision making, and created the need of jointly comparing a large number of treatments over multiple outcomes of interest.

A network meta-analysis (NMA) compares three or more treatments by using both direct comparisons of interventions within RCTs and indirect comparisons across trials based on a common comparator (Lumley 2002). By incorporating evidence from indirect comparisons, an NMA potentially includes more studies and can increase the precision of estimation. Statistical methods for univariate NMAs have been developed during the last twenty years (Lumley 2002; Caldwell et al. 2005; Lu and Ades 2006, 2009; Salanti et al. 2008; White et al. 2012). With the availability of multiple outcomes, recent literature advocates for jointly analyzing multiple outcomes in both conventional pairwise meta-analyses (Jackson et al. 2011; Riley et al. 2017) and NMAs (Efthimiou et al. 2014, 2015; Liu et al. 2018), which is advantageous over separated univariate analyses as it enables joint inferences on multiple outcomes, providing a more comprehensive evaluation of treatment options.

A major challenge of multivariate NMAs is modeling the correlation structure of different outcomes and treatment options. Using the random-effects model, the variance-covariance matrix among treatment comparisons and outcomes is assumed as a summation of the within- and between-study variance-covariance matrices. As the within-study variances are usually reported, in restricted cases this problem may be simplified when within-study correlations are also observed or can be calculated. For example, Jackson et al. (2018) considered the case when within-study correlations are reported by each study, and the unstructured between-study correlations are estimated through a method of moment approach. In studies where outcomes and exposures are all dichotomous and their contingency tables are reported, the within-study correlations may be calculated (Efthimiou et al. 2014). The within-study correlations among treatment comparisons for a specific outcome may be calculated if the standard errors of estimated effect sizes in all treatment groups are reported, taking advantage of the independence among the patients across different groups. However, the within-study correlations between two different outcomes are often not reported in RCTs (Jackson et al. 2011). Individual patient data (IPD) are needed to obtain such correlations, but they are hardly available in practice. To deal with the unknown within-study correlations, Efthimiou et al. (2015) adopted the method proposed by Riley et al. (2007), where synthesized correlation parameters were used to account for the marginal correlation between outcomes. Liu et al. (2018) focused on adjusting for outcome-reporting bias in multiple-outcome NMAs, and the unknown correlations were handled through copulas. In these methods, the effect sizes, heterogeneous variances, and correlations between outcomes need to be estimated. However, as more outcomes and treatments are considered simultaneously, the number of unknown parameters sharply increases, leading to new challenges in estimation of model parameters and valid statistical inference, as well as developing stable computational algorithms.

To address these issues, we propose a new framework for PAtient-centered treatment ranking via Large-scale Multivariate network meta-analysis (PALM). The contribution of our work is three-fold. First, we propose a parsimonious and robust modeling approach to address the unknown correlation structure that only requires specifying the marginal distribution of each outcome and treatment comparison. We further show that the proposed estimator is asymptotically consistent and normally distributed under mild regularity conditions. Second, we propose a computationally efficient algorithm which can generally achieve fast and stable convergence within a few iterations. Compared to the existing Bayesian methods (Efthimiou et al. 2015; Liu et al. 2018), the computational time is largely reduced. Third, we develop novel tools for patient-centered treatment ranking, including an intuitive star plot for presenting multivariate outcomes, as well as quantitative summaries accounting for both the point estimation and statistical uncertainty of the relative effectiveness of treatments.

The rest of the paper is organized as follows. Section 2 provides a motivating example of an NMA with more than a dozen treatment options along with five clinically important outcomes. Section 3 introduces the proposed method and a fast algorithm for estimation and statistical inference. In Section 4, we introduce multiple tools for personalized treatments ranking which take into account both the point estimates and the uncertainties estimated from our proposed method. In Section 5, we provide an in-depth NMA study evaluating methods of labor induction using our proposed method. Section 6 presents simulation studies to evaluate our method’s performance in terms of estimation bias, efficiency, and coverage probabilities. We conclude this article with a discussion in Section 7.

## 2 A Motivating Example: A Large-Scale Multivariate Network for Labor Induction Treatments

More than 20% of women undergoing an induction of labor each year in the United States (Centers for Disease Control and Prevention 2020). Cervical ripening is a necessary step prior to the beginning the induction itself. Many RCTs have evaluated various cervical ripening methods which include mechanical dilation of the cervix (e.g., with a Foley balloon catheter) and pharmacological interventions. Among the pharmacological methods, prostaglandins are the most commonly used both within and outside the U.S. (Boulvain et al. 2008; Alfirevic et al. 2015), with all forms of prostaglandins being shown to reduce the risk of cesarean when compared to no treatment or oxytocin alone (White et al. 2012). Among prostaglandin use, different formulations, dosing regimens, and routes of administration are used. In a recently published NMA, Alfirevic et al. (2015) compared 12 types of prostaglandin treatments with placebo and no treatment (14 treatment options in total), based on five clinically relevant outcomes: cesarean section, serious neonatal morbidity or perinatal death, serious maternal morbidity or death, vaginal delivery not achieved within 24 hours, and uterine hyperstimulation with fetal heart rate changes. Two hundred eighty RCTs comprising 48,068 women were selected from the Cochrane Pregnancy and Childbirth Group’s Database of Trials, where at least one of the five outcomes were reported in each trial.

Figure 1 shows the treatment networks of the five outcomes. Most trials reported only one or two outcomes, leading to substantial differences in the structures of the five networks. When considering five outcomes and 13 treatment comparisons (treating placebo as the reference treatment option) simultaneously, modeling the correlation structure is rather difficult. If assuming an unstructured correlation matrix, there are 1755 unknown parameters in the between-study covariance matrix^†^. Also, within-study correlation estimates were not reported in any individual study, creating difficulties for joint modeling multiple outcomes using the existing methods. In Alfirevic et al. (2015), three separate univariate NMAs were conducted one for each outcome of vaginal delivery not achieved within 24 hours, cesarean section, and uterine hyperstimulation with fetal heart rate changes, while the remaining two outcomes were not formally analyzed. As the decision making of labor induction treatments often involve multiple outcomes, this approach of separated NMAs is limited because it cannot provide a complete picture of the relative effectiveness of these treatment options with respect to the five relevant outcomes.

**Figure 1:**
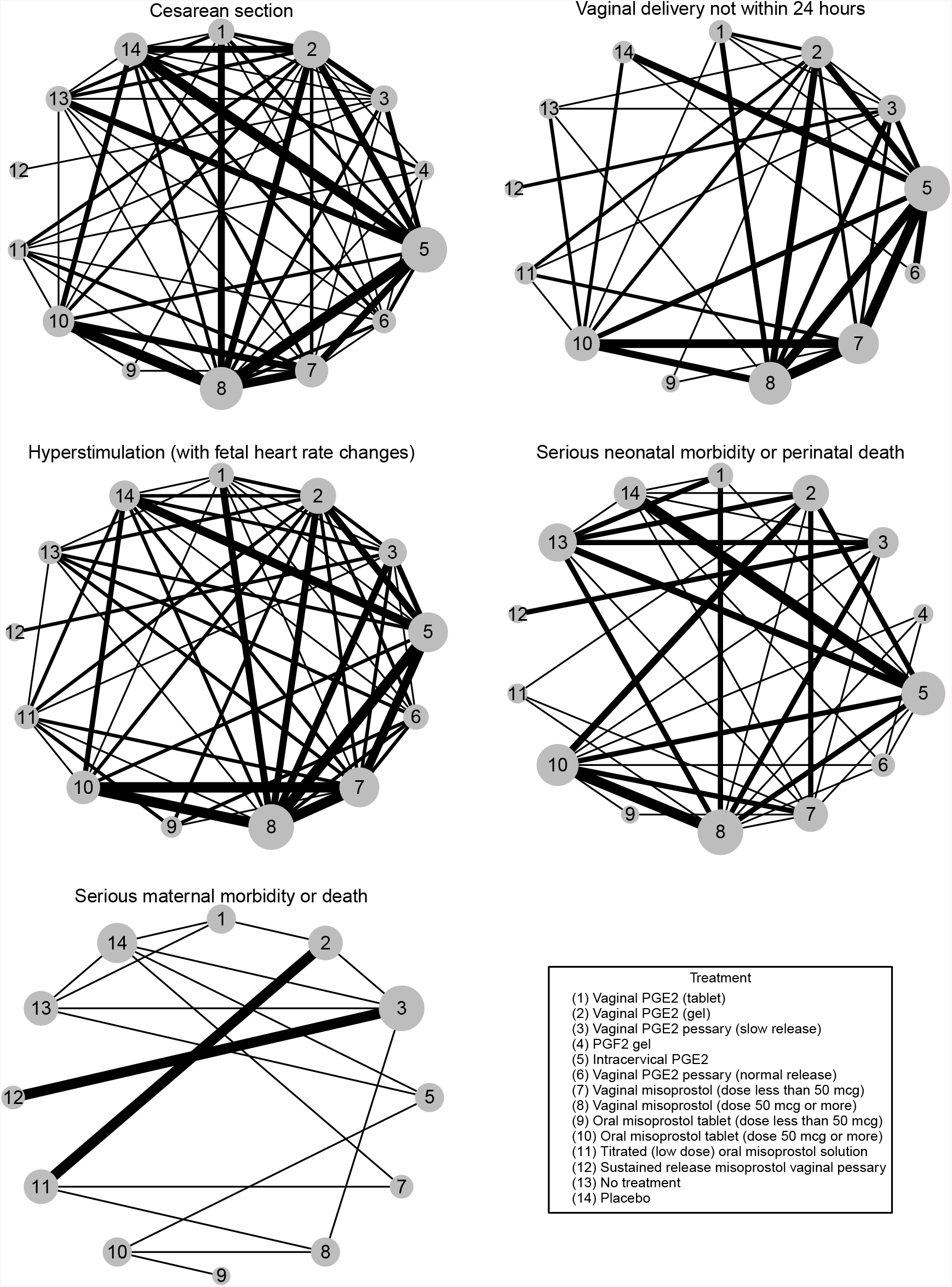
Network plots of the 14 treatment options and 5 outcomes in the network meta-analysis for prostaglandins in labor induction by Alfirevic et al. (2015).

These similar issues are shared in other NMAs with a large number of treatment options and multiple outcomes, which have become increasingly common across a wide spectrum of areas, including cancer (Mauri et al. 2008; Terasawa et al. 2013; Dulai et al. 2016), neuropsychiatric disorders (Cipriani et al. 2018; Slee et al. 2019; Pillinger et al. 2020; Bahji et al. 2020), cardiovascular diseases (Bash et al. 2012; Dunkley et al. 2012; Li et al. 2018), and among many others.

Motivated by the need of jointly analyzing networks comparing large number of treatments over multiple outcomes, as well as the need of new tools to support patient-centered clinical decision making, our methods are developed and introduced in the following sections.

## 3 Methods

### 3.1 Notation and model assumptions for multiple-outcome NMA

Suppose there are *n* studies in the network comparing a set of treatments *𝒯* = {A, B, C, …}. A set of *K* common outcomes, denoted by *{Y*_1_, *Y*_2_, …, *Y*_*K*_}, is of interest. Each study compares at least two treatments in *𝒯*. Suppose there are in total of *D* unique designs among all studies; denote them by *D* = {1, 2, …, *D*} and let *𝒯*_*d*_ be the subset of treatments compared in design *d*.

In this paper, we focus on a contrast-based framework. Let 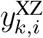 be the effect size estimate for outcome *k* from the *i*-th study comparing treatment *Z* to *X*, where *Z, X ∈ T*, and 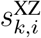 be the corresponding estimated standard error. For a study with design *d*, in practice, usually only estimates for |*𝒯*_*d*_| *−* 1 contrasts are reported, i.e., one treatment is set as the reference level, and all other treatments are compared to the reference.

To keep the notation simple and without loss of generality, in the following sections, we introduce our method using a three-treatment NMA (comparing A, B, and C) with two outcomes of interest, i.e., (*Y*_1_, *Y*_2_). We note that our algorithm is easily scale up to a large number of treatments or outcomes, as we will illustrate in Section 5. With treatments A, B and C, the possible designs include two-arm designs AB, BC, AC, and a three-arm design ABC. We denote 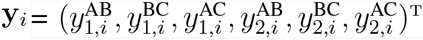, and 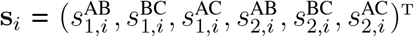for the estimated effect sizes and their estimated standard errors from the *i*-th study, respectively. Some studies may only report parts of the results while the remaining results are unobserved. Taking between-study heterogeneity into consideration, the random-effects model assumes that

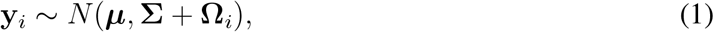

where 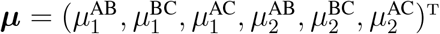 characterizes the true population effect sizes, and **Σ** is the between-study variance-covariance matrix,

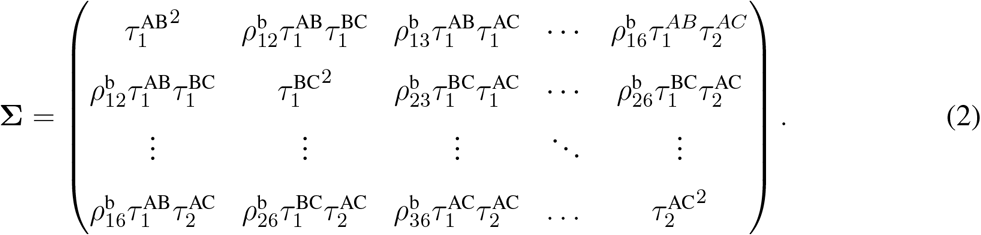

In the above matrix, 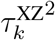 is the between-study variance of outcome *k* comparing treatment X and Z, for X, Y *∈ T*, and 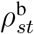 for 1 *≤ s < t ≤* 6 are the between-study correlations. We denote the between-study correlation matrix by **R**^b^ and the heterogeneity variances by 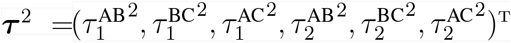. In addition, **Ω**_*i*_ is the within-study covariance of **y**_*i*_ :

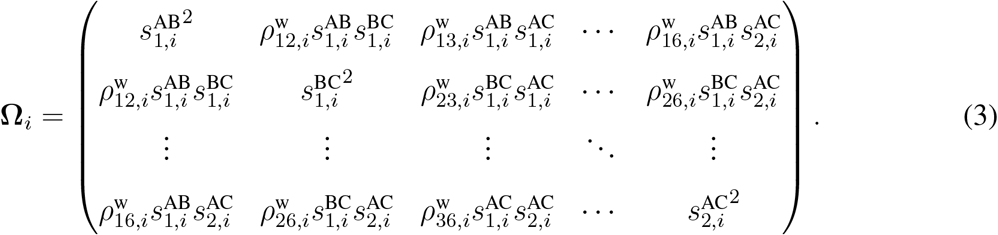

We denote the within-study correlation matrix by 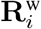. The elements of the within-study correlations 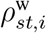 are rarely reported in published papers. One of the key motivations of our modeling approach, which is introduced in the next subsection, is to conduct valid statistical inference without the knowledge on the within-study correlation.

### 3.2 A robust and parsimonious modeling approach

One major challenge in fitting the model described in Equations (1)–(3) is the estimation of the large number of unknown parameters. Usually, the overall effect sizes ***µ*** and the heterogeneity variances ***τ*** ^2^ are parameters of primary interest, while the between-study correlation matrix **R**^b^ is often considered as nuisance parameters. For a large network, the dimension of **R**^b^ is large and estimating such a large number of parameters is challenging or infeasible.

We tackle this problem by constructing a pseudolikelihood likelihood function, adapting the idea of a composite likelihood which requires specifying only the marginal distribution of each 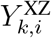, while avoiding the specification of the correlations between outcomes and contrasts. Essentially, it is done by assuming a working independent correlation structure for the multivariate outcomes. According to Lindsay (1988) and Cox and Reid (2004), this approach will obtain consistent estimates of the parameters of interest, and valid inference can be constructed using a sandwich-type variance-covariance estimator.

Different from White et al. (2012) and Efthimiou et al. (2014), for studies with a three-arm design ABC, we do not assume that all studies choose the same reference treatment. For example, if A is a common standard treatment or a placebo, in most cases 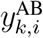 and 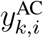 are reported for outcomes *k* = 1 and 2 with their standard errors 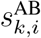 and 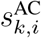. However, we do not eliminate the possibility that a study chooses B or C as the reference treatment. If we only observe 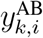 and 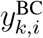 with 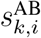 and 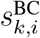, under the evidence consistency assumption, the treatment effect of C vs. A can be obtained as 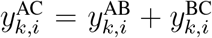. However, its standard error 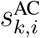 may not be obtained using the observed data 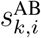 and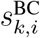, when the within-study correlation is not reported. Moreover, if effect sizes and standard errors for all comparisons of AB, BC, and AC are reported, we suggest incorporating all of them in the analysis, instead of only using those of AB and AC. Although 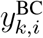 can be derived from 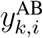 and 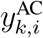, its information is not redundant because both 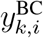 and 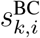 contribute to the estimation of 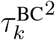 and thus improve the estimation of other parameters. More importantly, by simultaneously incorporating all reported outcomes, we can avoid the issue that estimation results may depend on the choice of the reference level.

To construct the pseudolikelihood, let 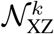 be the subset of studies which report effect sizes and standard errors of outcome *k* for comparing treatments X and Z. Also, let the sample size be *n*_XZ,*k*_, where XZ *∈* {AB, BC, AC}. The log-pseudolikelihood function can be written as

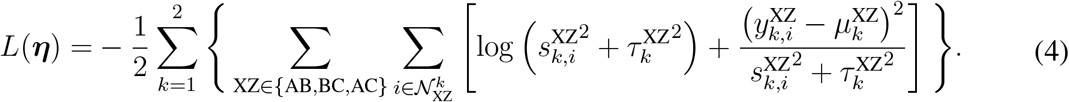

Under the evidence consistency assumption, the parameters satisfy the constraint

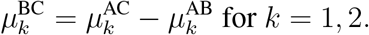

Therefore, we only need to estimate the parameters 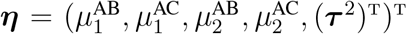. By maxi-mizing the log-pseudolikelihood function in Equation (4), we obtain the estimated parameters:

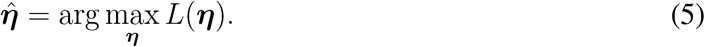

The following theorem describes their limiting distribution.

#### Theorem 1

*Under Assumptions C1-C5 in Appendix F of the Supplementary Material, the maximum pseudolikelihood estimator defined in Equation (5) satisfies*

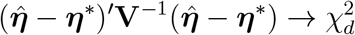

*as the total number of trials n increases, and the variance-covariance structure* **V** *is defined as* **V** = **I**^*−*1^**ΛI**^*−*1^, *with*

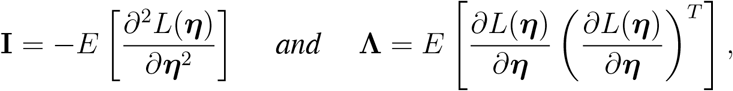

*and* ***η***^*∗*^ *denotes the true value of parameter* ***η***.

A sketch of the proof is provided in Appendix F of the Supplementary Material.

#### Remark 1

(Asymptotic normality and implication to univariate NMA) Theorem 1 shows the maximum pseudolikelihood estimator defined in Equation (5) is asymptotically normal. It is a general conclusion regardless of the true underlying correlation structure of the observations. Our method can also be applied to univariate NMA with a straightforward adjustment to the pseudolikelihood function in Equation (4). In practice, the restricted maximum likelihood (REML) approach can be adapted to improve the finite-sample estimation of the components in the variance-covariance matrix by adding a term 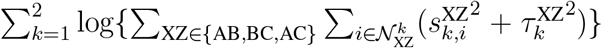 in Equation (4), and the REML estimator shares the same asymptotic distribution as the maximum likelihood estimator defined in Equation (5).

#### Remark 2

(Information orthogonality) The information matrix **I** is a block-diagonal matrix. With a slight abuse of notation, let 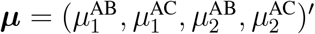. Denote

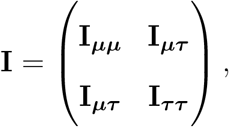

where **I**_***µµ***_ = *E*[*∂*^2^*L*(***η***)*/∂****µ***^2^], **I**_***ττ***_ = *E*[*∂*^2^*L*(***η***)*/∂*(***τ*** ^2^)^2^] and **I**_***µτ***_ = *E*[*∂*^2^*L*(***η***)*/∂****µ****∂*(***τ*** ^2^)]. It can be shown that **I**_***µτ***_ = **0**, implying that ***τ*** ^2^ and ***µ*** are considered as information orthogonal (Liang et al. 1995). This information orthogonality is stronger than other types of weak orthogonality conditions defined in Liang (1983) and Liang et al. (1995), and it provides the following simpler form when making inference on ***µ***. The variance-covariance matrix of 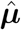 is simplified as

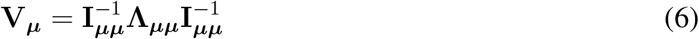

where

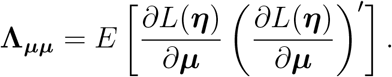

We can estimate **V**_***µ***_ empirically using 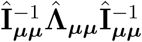, where Î_***µµ***_ and 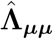 are the corresponding submatrices of the sample-version matrices Î and 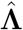, defined in Appendix A of the Supplementary Material. The information orthogonality provides a simplified way of estimating the variance-covariance matrix of the estimated effect sizes 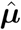, which only involves submatrices of the information matrices. In addition, the information orthogonality reveals the minimal impact of estimation of the between-study variance ***τ*** ^2^ on the estimation of the effect sizes ***µ***. In the case of having a relatively small sample size, the variance term ***τ*** ^2^ is often hard to be estimated accurately. With the property of the information orthogonality, the estimation of ***µ*** is not sensitive to the estimation of ***τ*** ^2^ (Liang 1983).

### 3.3 A fast iterative algorithm for parameter estimation

We propose an iterative algorithm for estimating the model parameters by maximizing the log-pseudolikelihood function in Equation (4) based on the following observations. First, this function can be written as the summation of two separate parts, i.e., 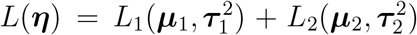, where 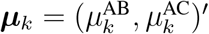, and 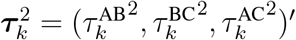 for *k* = 1 and 2, with

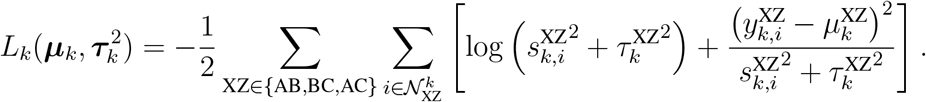

This implies that the estimation of ***η*** can be done by estimating 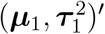 and 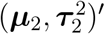 separately. Second, if 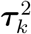 is fixed at some value 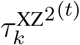, the parameter ***µ***_***k***_ can be estimated by solving a system of linear equations. More specifically, denote

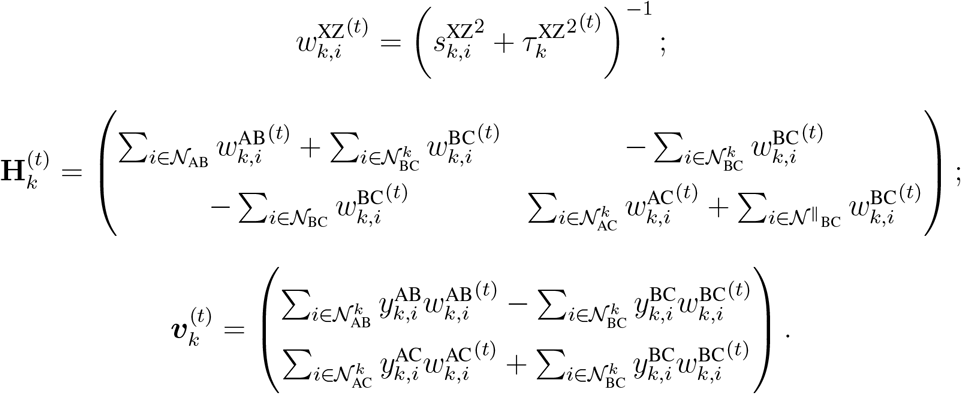

Maximizing 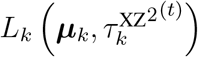 over ***µ***_*k*_ yields

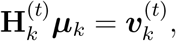

where ***µ***_*k*_ is the solution of the above system of linear equations. Thus, we propose to decompose the optimization into iteratively maximizing over ***µ***_*k*_ and 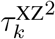. The detailed steps are summarized in Algorithm 1.

The initial value for 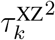 can be obtained by fitting a separate univariate random-effects meta-analysis for outcome *k* and comparison XZ. Based on our numerical study, the initial value can be chosen from a wide range of values, yet the results are not sensitive to the choice of initial values. The quantity *δ* is a specified tolerance level for convergence which is set to 10^*−*6^ in this paper. In each iteration, updating ***µ*** by solving the linear system equations is computationally efficient. Based on our simulations and application, the convergence is usually achieved within five iterations.

#### Algorithm 1 Iterative estimation

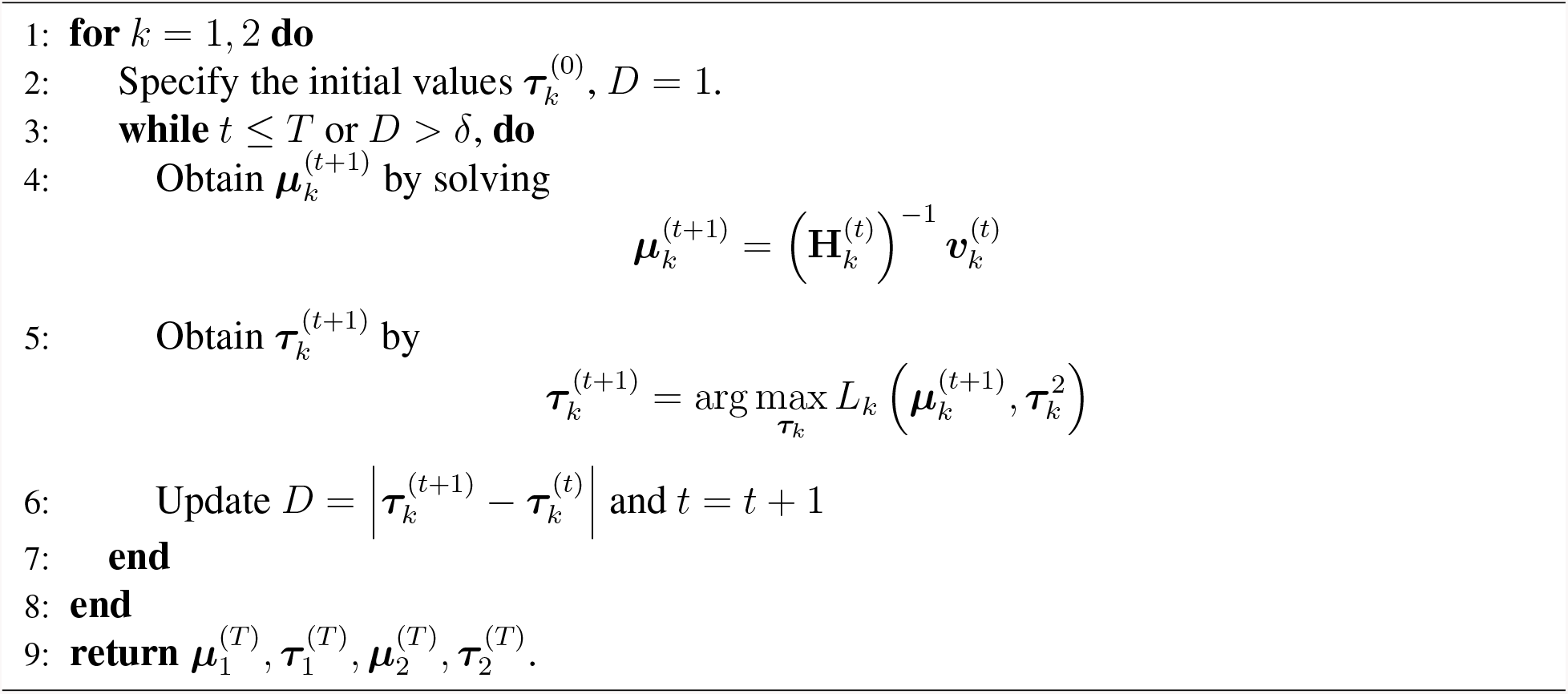

#### Remark 3.

(Special case with equal between-study variances) As suggested in White et al. (2012) and Efthimiou et al. (2015), we might want to reduce the model complexity and assume all treatment comparisons have the same between-study variance 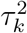 for each outcome *k* = 1, 2. In this case, the log-pseudolikelihood in Equation (4) reduces to

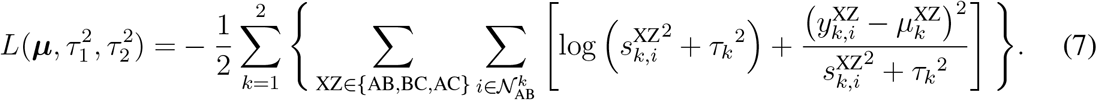

The optimization algorithm is modified correspondingly by mapping 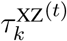 to 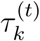 in **H**^(*𝒯*)^ and ***v***^(*𝒯*)^. The detailed algorithm and a variance estimator 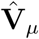 provided in Appendix B of the Supplementary Material. We noted that due to information orthogonality, the inference on the population effect sizes ***µ*** is not affected by the potential violation of the equal between-study variance assumption.

### 3.4 Inconsistency model

The foregoing methods are based on the assumption of evidence consistency; that is, 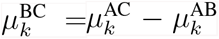 for each trio of treatment pairs AB, AC, and BC and for each outcome *k*. In practice, this assumption may be violated for some NMAs, and models that permit evidence inconsistency may be considered. The inconsistency may arise from various factors, including outlying, low-quality, or high-risk-of-bias studies (Guyatt et al. 2011). In the presence of potential evidence inconsistency, we may apply both inconsistency and consistency models to the NMA dataset, and compare their corresponding goodness of fit, measures of model selection (e.g., Akaike information criterion), and estimated heterogeneity extents. If the inconsistency is found to be substantial and meaningful, their causes should be explored; otherwise, great cautions should be employed in the interpretation of results that are affected by unexplained inconsistency.

Various statistical methods are available to adjust or test for evidence inconsistency (Lu and Ades 2006; Salanti 2012; Chaimani et al. 2013); for example, Dias et al. (2013) suggested the unrelated mean effects (UME) model, which treats 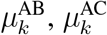, and 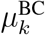 as three separate, unrelated parameters to be estimated. It is straightforward to extend the proposed method to the setting of evidence inconsistency. Specifically, the log-pseudolikelihood function in Equation (4) remains the same; however, without using the consistency equation, the parameters ***η*** to be estimated additionally include 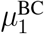 and 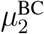. All theoretical results and algorithms can be readily modified by including these additional parameters. We provide detailed model formulation and algorithm in Appendix C of the Supplementary Material.

## 4 A Framework for Personalized Treatment Ranking

### 4.1 Star plot: a novel visualization tool for multiple outcomes

To jointly visualize the treatment efficacy among all the outcomes, we propose a visualization tool, termed as the *star plot*, which is inspired by the radar chart, in which multivariate outcomes are represented as multiple axes in two dimensional plot. See Figure 2 as an illustration. For a particular treatment, the radar chart presents the impacts of this treatment on all five outcomes as dots on five axes that are arranged radially around a common origin. For each outcome the origin of the axis represents the value of the lowest ranking (i.e., the worst treatment) effect estimate for the corresponding outcome among all treatments. In contrast, the utmost point represents the effect size of the highest ranking (i.e., the best) treatment from among all treatments. Lines are added to connect dots to obtain a polygon, and the area of the polygon is often used to quantify the overall treatment impact considering all the outcomes, with the larger area indicating better overall performance (Zhu et al. 2017). The advantage of the radar plot is that it presents both the individual outcomes and also provides an intuitive way of summarizing the overall comparison based on all outcomes. However, the area of the polygon is sensitive to the order of the outcomes in the plot. We provided an example in Appendix D of the Supplementary Material to show that changing the order of the outcomes can lead to different ranking of the treatments in a radar plot.

**Figure 2:**
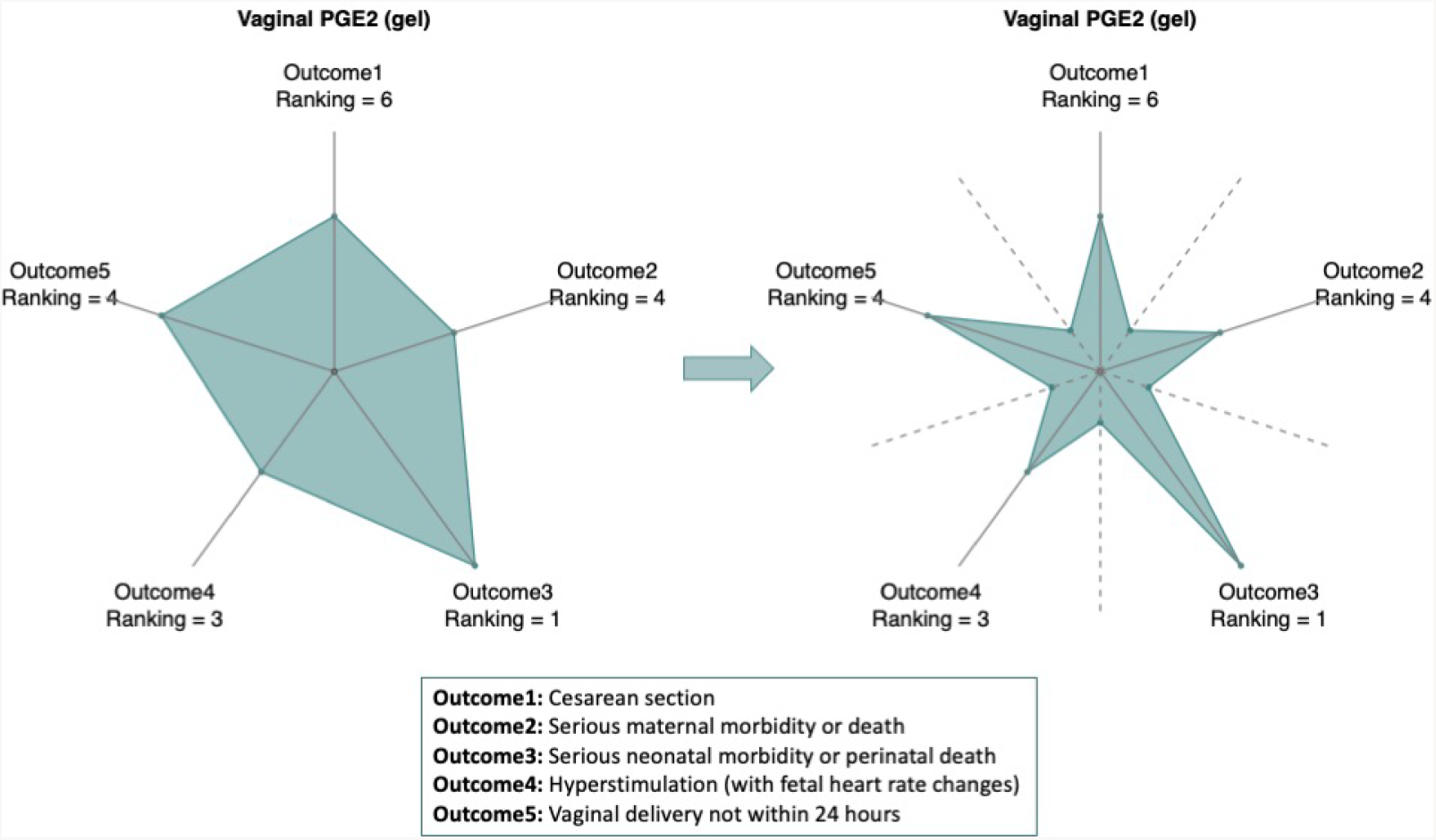
An illustration of the construction of the star plot (on the right), which is based on the radar chart (on the left). The radar chart displays multiple outcomes using dots on multiple axes which are arranged radially with a common origin on a two-dimensional plot. To construct the star plot, we keep the main axes of the radar plot (on the left) and add five auxiliary axes (the dashed lines) each in between of a pair of two adjacent main axes. Dots with equal distance to the origin are place auxiliary axes. We connect the dots in order and obtain a star shape region. The ranking of a treatment based on each individual outcome is placed next to the corresponding axis, and the area of the shaded region is used as an overall quantification of the treatment based on all outcomes.

To address this limitation, we propose a new visualization tool which provides treatment ranking invariant to how the outcomes are ordered in the plot. As shown in the right panel of Figure 2, in addition to the five main axes (the solid lines) which present the individual outcomes, we propose to add an auxiliary line (dashed line) between each pair of two adjacent main axes (solid line). In our example, for a given treatment, we place five dots on the main axes to represent each outcome and five dots each on an auxiliary line. These five auxiliary points equally distant from the center, however, the distance can be arbitrary. If we choose a distance near the origin and connect all dots in order, we can obtain a shaded region for each treatment which has a star like shape, hence the name *star plot*. Each angle of a star represents one outcome where longer length indicates better performance of this outcome for the specific treatment. In the star plot, we also indicate next to the angle the ranking of the treatment for the corresponding outcome. Importantly, the area of the shaded region, which is now invariant to the ordering of the outcomes, provides an overall quantification of the treatment impact across all the outcomes where a larger area indicates better overall performance of the treatment when taking all outcomes into consideration.

Using the star plot, we can compare treatments based on any subsets of the individual outcomes, as well as considering all the outcomes jointly. In addition, the star plot can be customized to include only outcomes of interest, and/or can assign different weights to different outcomes depending on their importance to individual patients. For example, our clinical expert suggested that the two outcomes, vaginal delivery not achieved within 24 hours and uterine hyperstimulation, may not be as important as the other three outcomes in terms of clinical decision making. In this case, the star plot can be modified (see Figure S9 in the Supplementary Material) to assign less weights to these two outcomes.

### 4.2 A weighted utility function

For univariate NMAs, several quantitative measures have been developed for ranking interventions. For example, the *Pbest* calculates the probability of each treatment to produce the best outcome; a *rankogram* is a graphical presentation of the probabilities for any intervention having specific ranks (Salanti et al. 2011). More specifically, the probabilities of *m* treatments ranking at *m* places using a combined histogram, where the *i*-th bar is partitioned into *m* parts with each part denotes the probability of a treatment ranking at the *i*-th places. In addition, the surface under the cumulative ranking curve (SUCRA) is commonly used as a quantitative way to rank interventions (Salanti et al. 2011; Trinquart et al. 2016). When considering more than one outcome, Mavridis et al. (2019) recently proposed to calculate the probability of all outcomes of treatment *Z* being better than the corresponding outcomes of treatment *X* plus some prespecified clinical important values, i.e., 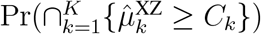.

In this paper, we proposed to rank the treatment based on a joint consideration of multiple outcomes, through a personalized utility function which incorporates individual considerations on different outcomes. For example, when evaluating treatment options for labor induction, risk for cesarean section is often an important efficacy measure as cesarean section has been associated with many immediate and long-term complications (Silver 2012; Caughey et al. 2014). On the other hand, treatment options with higher risk for serious adverse events such as maternal or neonatal morbidity or death would not be preferred. In addition, some patients may care more about whether virginal delivery can be made within 24 hours compared to others. Therefore, these outcomes need to be balanced against one another when evaluating induction methods.

Without loss of generality, suppose outcome 1 measures the treatment efficacy and outcome 2 measures safety. To jointly consider the two outcomes, we define a combined utility function by assigning different weights to the two outcomes and calculate the weighted sum. Let *w*_1_ and *w*_2_ be the weights for outcomes 1 and 2, respectively. To compare two treatments X and Z, we define the weighted utility function as

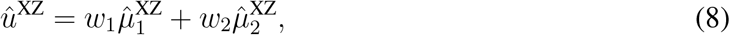

and the treatment ranking is based on the weighted utility function. The weights can be specified by clinicians, patients or stakeholders when they make decisions on choosing the best treatment. This framework can be easily extended to utility functions of more than two outcomes.

### 4.3 Pairwise comparison of two treatments based on the utility function

For pairwise comparison, we are interested in the quantity 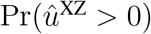, which measures the probability of treatment X being better than Z, under a certain weight assignment. From Section 3.2, we can approximate the joint distribution of ***û*** by a multivariate normal distribution with mean ***µ*** and variance 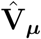 defined in Remark 2.

When X is the reference drug A, then we define ***e***_AZ_ as an indicator vector with the same length as ***µ***_1_, where the *i*-th entry of ***e***_AZ_ indicates whether the *i*-th entry of ***µ***_1_ involves the contrast AZ. For example, in the three-arm setting used in the previous sections, we define 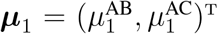; thus, we have ***e***_AB_ = (1, 0)^T^, and ***e***_AC_ = (0, 1)^T^.

If neither *X* nor *Y* is the reference drug, we have

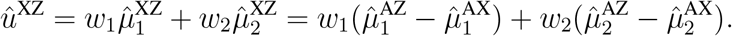

We then define ***e***_XZ_ as the indicator vector with the same length as ***µ***_1_, where the *i*-th entry of ***e***_XZ_ is *−*1 if the *i*-th entry of ***µ***_1_ involves the contrast AZ and is 1 if that of ***µ***_1_ involves the contrast AX. For example, in the three-arm case, we have ***e***_BC_ = (*−*1, 1)^T^. Let ***w***_XZ_ = (*w*_1_***e***_XZ_, *w*_2_***e***_XZ_)^T^. By the property of multivariate normal distribution, we provide an estimator 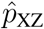 for the quantity 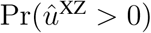, calculated as

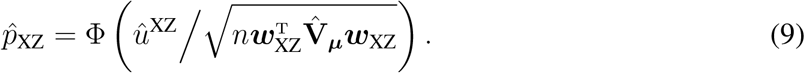

### 4.4 Treatment ranking of more than two treatments using weighted SUCRA

When more than two treatments are to be compared, we are interested in estimating the probabilities for any treatment assuming any possible rank. For example, consider there are *m* treatments of interest, assume them to be *{X*_1_, …, *X*_*m*_}. We define *q*_*ij*_ to be the probability of treatment *X*_*i*_ ranking at the *j*-th place based on the defined utility function in Equation (8). To estimate *q*_*ij*_, we first define ***e***_*i*_ to be an indicator function of the *i*-th treatment where the *i*-th entry of ***e***_*i*_ indicates whether the *i*-th entry of ***µ***_1_ involves the contrast *AX*_*i*_. If *X*_*i*_ = *A*, i.e., the reference treatment, we have ***e***_*i*_ = **0**. In this way, we obtain a design matrix ***D*** = (***e***_1_, …, ***e***_*m*_)^T^, and

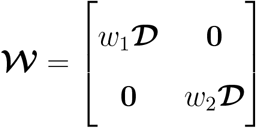

and we propose to estimate *q*_*ij*_ using the following sampling method.

#### Algorithm 2

Estimating *q*_*ij*_ via random sampling

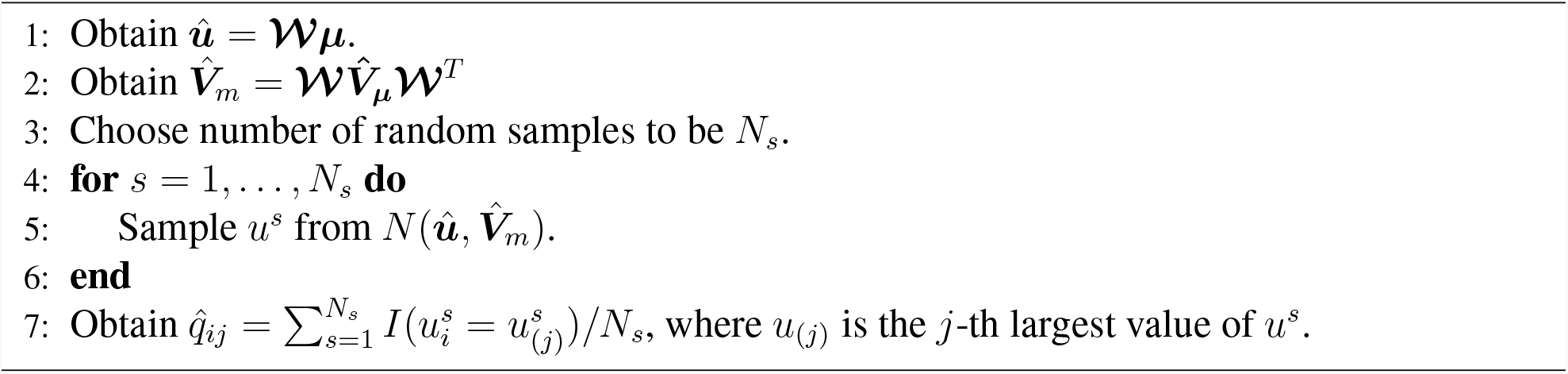

With the estimated 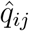 we define the weighted SUCRA as

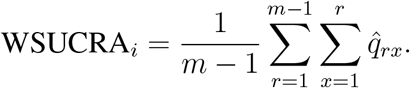

This WSCURA synthesizes multiple treatment effects into a single value, which ranges from 0 to 1. Higher the value of the WSUCRA indicates a better treatment based on the personalized utility function; WSUCRA= 0 indicates an always worst treatment, and WSUCRA= 1 indicates the best ranked treatment, making it easy for both interpretation and decision making.

#### Remark 4

In addition to SUCRA, we can also define the weighted P-score as

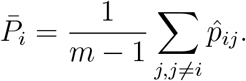

It can be shown that P-score is numerically nearly identical as SUCRA, which provides a way to estimate SUCRA without resampling by directly using Equation (9).

## 5 Multivairate NMA of labor induction methods

We applied our method to the dataset from the NMA for labor induction treatments introduced in Section 2. Choosing the placebo as the reference treatment, we jointly modeled all five outcomes using the network shown in Figure 1. As suggested by the simulation study in Section 6, assuming the same between study variance across different treatment comparisons can further reduce the model complexity without creating additional bias on estimation of treatment effects. Thus, we used the likelihood function defined in Equation (7) with a REML correction. The algorithm took less than 3 seconds to converge on a computer cluster node (HP DL165-G6) with AMD 2.2 GHz CPU and 16GB memory. We presented the detailed estimation results in Appendix E of the Supplementary Material, where Figure S2 shows the forest plots of the estimated log odds ratios and the corresponding 95% confidence intervals for the 13 treatment options compared to the placebo across the five outcomes, and Figure S3 presents the pairwise comparisons of any two treatments over the five outcomes.

Among the 12 prostaglandin treatment options, 8 are commonly used both within and outside the United States (Boulvain et al. 2008; Alfirevic et al. 2015). They are vaginal prostaglandin E2 (PGE2) gel, vaginal PGE2 pessary (slow release), intracervical PGE2, vaginal misoprostol (dose *<*50 *µ*g), vaginal misoprostol (dose *≥* 50 *µ*g), oral misoprostol tablet (dose *<* 50 *µ*g), oral misoprostol tablet (dose *≥* to 50 *µ*g), and low dose oral misoprostol solution. The four other groups are not currently manufactured for routine use (vaginal PGE2 tablet, PGF2 gel, vaginal PGE2 pessary (normal release), and sustained release misoprostol vaginal pessary). Therefore, we focused on comparisons of the former 8 treatments to the placebo in the following.

Jointly analyzing five outcomes using the proposed framework has led to a few new insights. First, we are able to intuitively visualize the treatment comparisons based on all five outcomes (See Figure 3). As we introduced in Section 4.1, each star of the plot represents a treatment and each angle of the star corresponds to one of the five outcomes. The longer length of the angle of the star, the better the treatment performed on the corresponding outcome. Starting at the top and moving clockwise we observed that low dose misoprostol solution is ranked as number 1 as it had the lowest risk for cesarean section (star #4), intracervical PGE2 had the lowest risk for serious maternal morbidity (star #1), vaginal PGE2 (gel) was the best treatment based on the risk for serious neonatal morbidity or perinatal death (star #7), low dose oral misoprostol tablet had the lowest risk for hyperstimulation (star #3), and high dose vaginal misoprostol had the highest chance for vaginal delivery within 24 hours (star #5).

**Figure 3:**
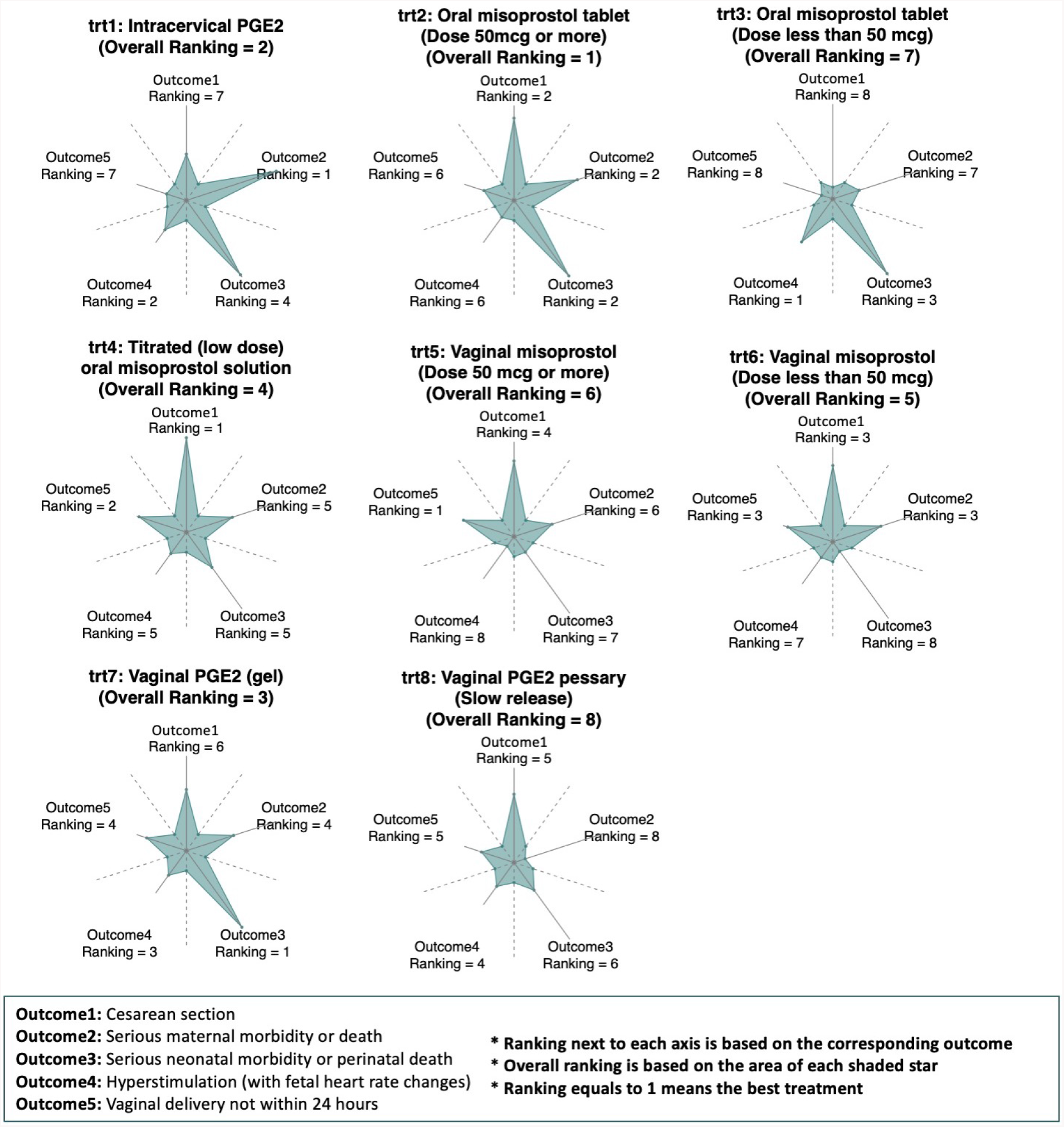
A star plot to compare 8 treatment options for labor induction across five outcomes. The distance of the dots on the main axes to the common origin are determined by the estimated effect sizes of the corresponding treatment from the proposed method. The distance of the dots on the auxiliary axes are set to 1*/*5 of the total length of the axis. The ranking of a treatment based on each individual outcome is placed next to the corresponding axis. The overall treatment ranking based on the area of the shaded region is indicated in the plot.

The overall treatment rankings based on all 5 outcomes (ordered by the area of the stars) were also indicated in the plot, and it showed vaginal PGE2 (gel) had the best overall effect among all 8 treatment options, while vaginal PGE2 pessary (slow release) ranked the lowest. Again, the star plot can be tailored to up-weight or down-weight certain outcomes (See Figure S9 in the Supplementary Material for an example).

Secondly, we considered the risk for cesarean section and serious neonatal morbidity or perinatal death combined. These two outcomes are of great clinical importance when evaluating efficacy and safety. The efficacy outcome, cesarean section, has been associated with many immediate and long-term complications with a strong national push to lower the rate of first-time cesarean sections (Silver 2012; Caughey et al. 2014). Determining which cervical ripening/induction methods are more effective at lowering the cesarean rate would have huge public health implications. The neonatal outcome of morbidity or death is of critical importance as no clinician or patient would choose an effective method if it caused harm to the neonate. Therefore, these two outcomes need to be balanced against one another and incorporated as major outcomes in studies evaluating induction methods.

Figure 4 presents the estimated effect sizes of the 8 treatments and their corresponding confidence intervals using a two dimensional-scatter plot. The drugs with lower risk of cesarean section and serious neonatal morbidity or perinatal death are in the upper right panel of the scatter plot. Most of the 8 treatments had significantly lower risk for cesarean section with the exception of low dose of oral misoprostol tablet, and intracervical PGE2 (marginally significant). For neonatal morbidity or perinatal death, however, no treatment was observed to be significantly better than the placebo. Based on the point estimates (dots in the plot), the high dose oral misoprostol tablet had low risk for both cesarean section and neonatal or perinatal death. Titrated oral misoprostol solution has the lowest risk for cesarean section but the risk for neonatal morbidity or perinatal death is relatively high. Vaginal PGE2 gel and intracervical PGE2 have similar risk for neonatal morbidity or perinatal death as the high dose oral misoprostol tablet, but their risk for cesarean section are higher, although the confidence intervals overlap. Given the different pharmacokinetics of prostaglandins based on the route of administration (Tang et al. 2002; Bygdeman 2003; Khan et al. 2004), it is plausible that higher doses or vaginal misoprostol would not have the same risks when administered orally. This NMA suggests that oral misoprostol with doses *≥*50 *µ*g is superior to other preparations and dosing regimens.

**Figure 4:**
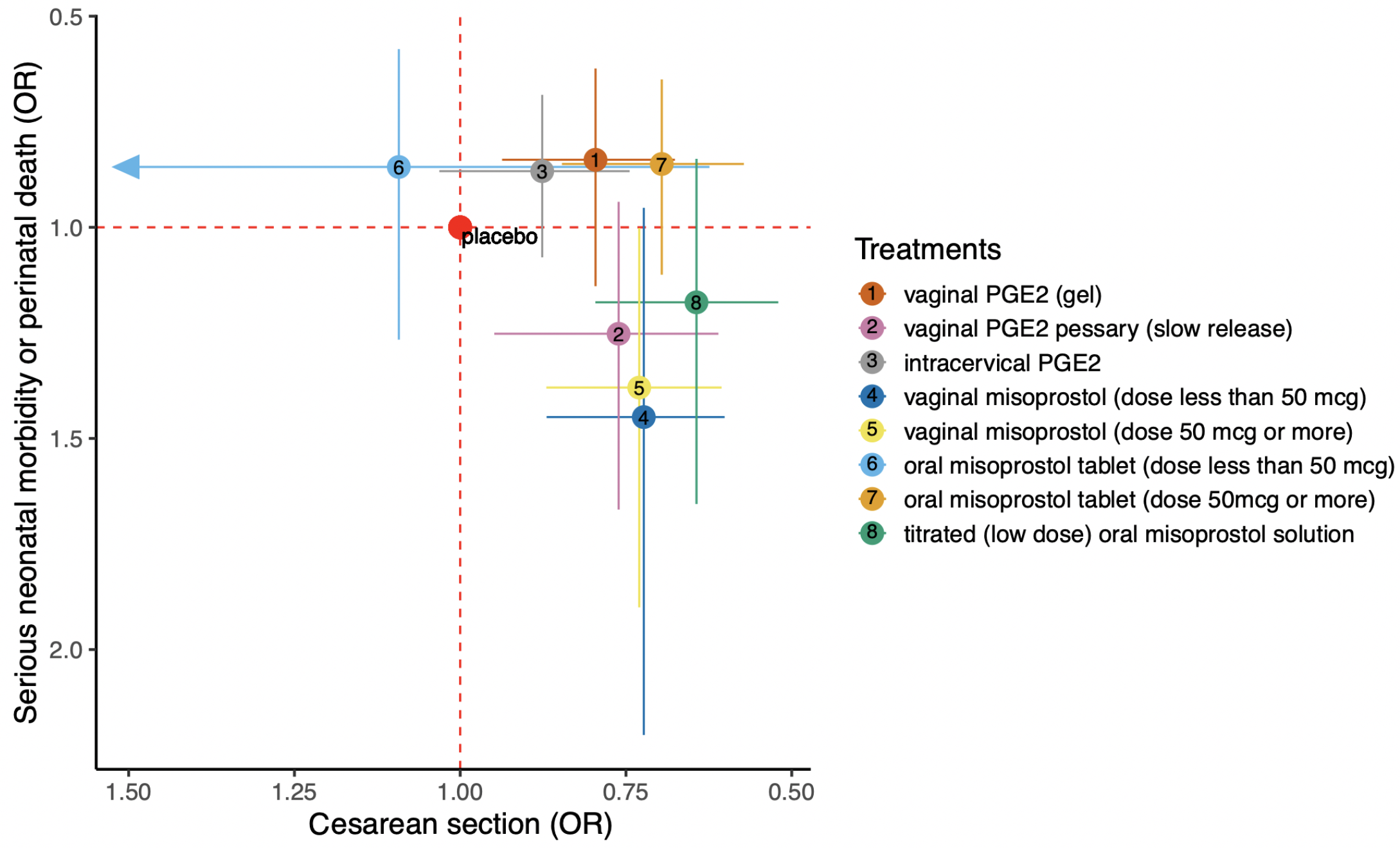
The estimated odds ratios of cesarean section and serious neonatal or perinatal death, with the corresponding 95% confidence intervals for the 8 treatment options for labor induction.

Thirdly, we compared the 8 treatments by considering cesarean section and maternal morbidity jointly, as both patients and clinicians usually feel that cesarean section and maternal morbidity are two competing risks. We defined a utility function as a weight sum of these two outcomes, and calculated the probabilities of each treatment ranking from the best to the worst based on the utility function. Figure 5 shows the rankogram for the eight treatment options based on the personalized utility function. For example, when a pregnant woman or physician weighs risk of cesarean section higher than the risk of maternal morbidity (Figure 5 panel 1: 90% weight to cesarean and 10% weight for maternal morbidity), titrated oral misoprostol solution was more likely to be better than other treatment options, while oral misoprostol tablet (low dose) was more likely to be the worst option. When a woman or physician weighs the two outcomes equally (panel 2: 50% weight for cesarean and 50% weight for maternal morbidity), oral misoprostol tablet (low dose) has the most probability to be the best option, although titrated oral misoprostol solution was ranked very closed to it. The oral misoprostol tablet (low dose) was rank as the worse option. If a woman or physician weighs maternal safety and morbidity more important than the risk of cesarean (panel 3: 90% weight for maternal morbidity and 10% weight for cesarean), the intracervical PGE2 was more likely to be better than other options while vaginal PGE2 pessary (slow release) would not be recommended. The WSUCRA of the eight treatment options gave a more clear ranking of the 8 treatment options, and we included the results in Appendix E of the Supplementary Material. We also explored other utility functions of the five outcomes, and the results are shown in Appendix E of the Supplementary Material.

**Figure 5:**
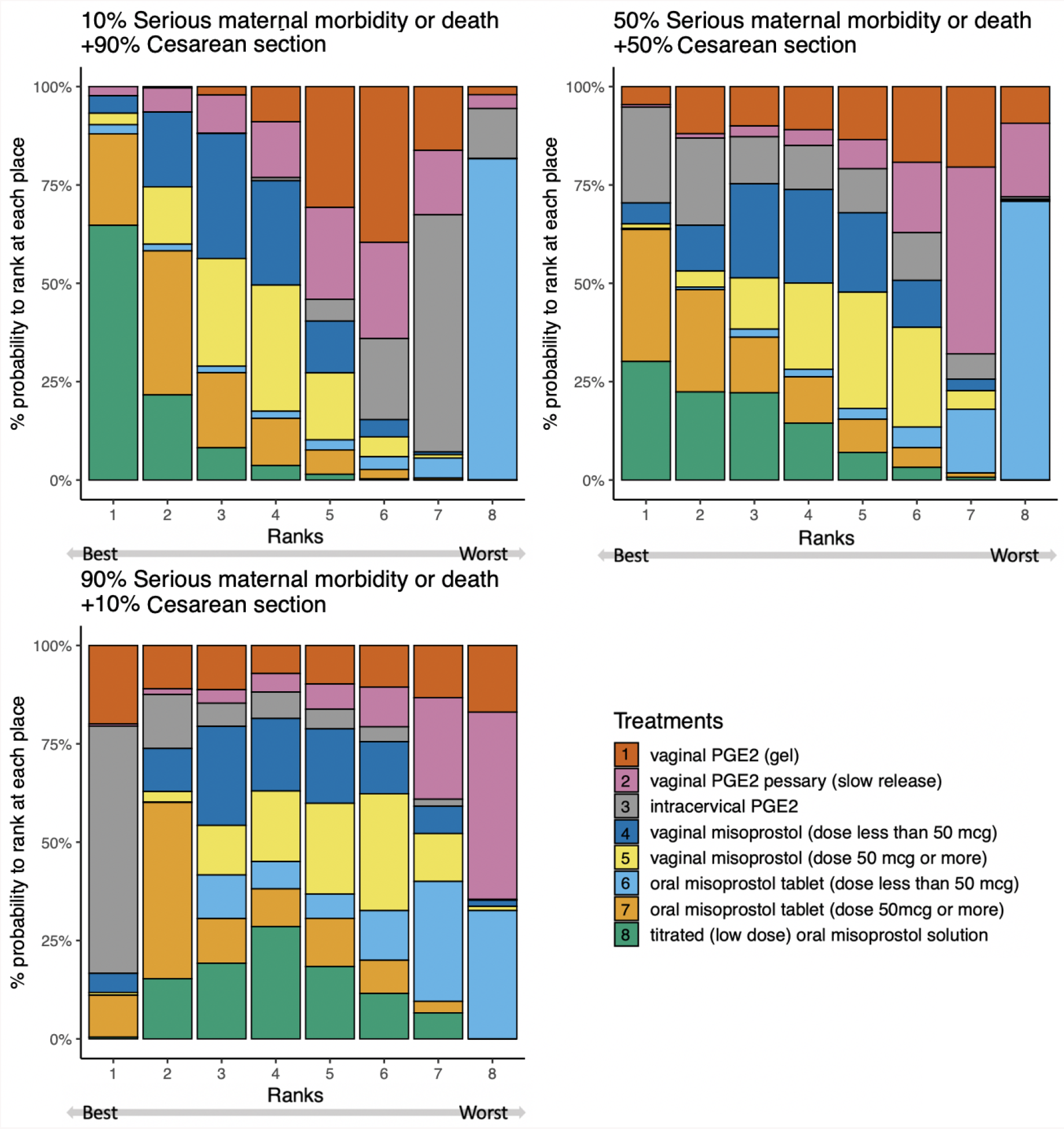
Ranking of the 8 treatment options for labor induction regarding three different weight options for cesarean section and maternal morbidity. In each plot, the *j*-th bar represents the probabilities of each treatment ranking in the *j*-th place.

Finally, in addition to the above analyses, we performed a sensitivity analysis by removing studies that were evaluated with high risk of bias in Alfirevic et al. (2015). The sensitivity analysis included 162 studies out of the 280 studies, and the results were mostly consistent with the analysis including all studies, which can be found in Appendix E of the Supplementary Material.

In summary, this investigation of treatment options for labor induction using our method and visualization tools has provided a comprehensive picture of the labor induction treatment comparisons based on five clinical relevant outcomes, and offers a patient-centered treatment ranking approach to better support clinical decision making.

## 6 Simulation Study

We conducted simulation study to evaluate the performance of the proposed methods. Without loss of generality, we considered three-arm designs with two outcomes and use the contrast-based data generating mechanism. Let 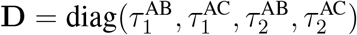, and we first generated

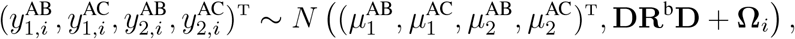

where **R**^b^ is the between-study correlation matrix and **Ω**_*i*_ is the within-study variance. Also, we set 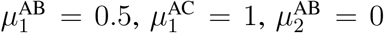, and 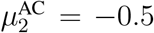. By the evidence consistency assumption, 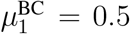 and 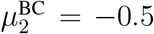. Then, we obtained 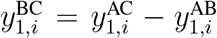 and 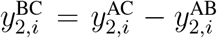. The between-study variance 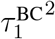 and 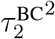 can be calculated as

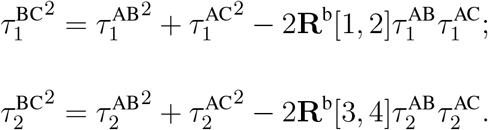

We considered the following two settings. In setting 1, between-study variances were equal for each outcome, i.e., 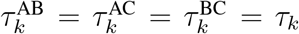. We set *τ*_1_ = 0.5 and *τ*_2_ = 0.6. In this setting, **R**^b^[1, 2] = **R**^b^[3, 4] = 0.5, and we set the between-study correlation matrix to

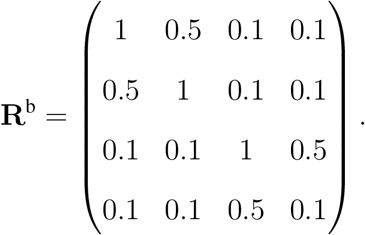

In setting 2, between-study variances were not equal. We set 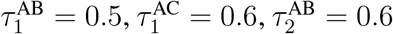, and 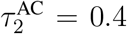. We set the between-study correlation matrix **R**^b^ to have an exchangeable correlation structure with correlation coefficient 0.1; that is, all its off-diagonal elements were 0.1. Therefore, we obtained that 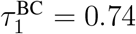 and 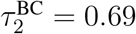.

Under each scenario, we randomly generated **Ω**_*i*_ for each observation and consider the sample size for each design were equal; that is, *n*_AB_ = *n*_BC_ = *n*_AC_ = *n*_ABC_ = *n*. We compared the proposed method under two assumptions, i.e., equal between-study variance and unequal between-study variances, corresponding to Algorithm 1 (PALM) introduced in Section 3.3 and Algorithm S1 (PALM-equal) from Remark 3 and the Supplementary Material. To compare the two algorithms, we calculated the bias, model-based variance, empirical variance, and coverage probability based on 1000 simulation replicates.

Tables 1 and 2 present the simulation results under simulation settings 1 and 2, respectively. When the between-study variances were the same for different treatment comparisons (see Table 1), the algorithm assuming equal variance (PALM-equal) had smaller bias, and slightly underestimated variance, which led to coverage probabilities slightly below 95%. The method assuming unequal between-study variances (PALM) had much larger bias when sample size is small, compared to PALM-equal. The variance was also larger than PALM-equal, with the relative efficiency ranging from 1.5 to 2. PALM had better coverage probabilities, which ranged from 93.2% to 95.8%. When the between-study variances are unequal (see Figure 2), PALM-equal still had smaller bias compared to PALM. PALM-equal had smaller coverage probabilities, especially when sample size is small. When sample size was large, the coverage probabilities are around 92.0%. PALM had slightly overestimated variances, and coverage probabilities ranged from 93.6% to 96.6%.

**Table 1:** Comparison of the bias, model based standard error (MBSE), empirical standard error (ESE) and the coverage probability (CP) of the effect sizes of treatment comparison AB and AC estimated from the PALM and PALM-equal methods, under the Setting 1 where 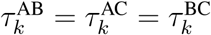 for *k* = 1, 2.

| No. of studies | Method | Comparison AB |  |  |  | Comparison AC |  |  |  |
| --- | --- | --- | --- | --- | --- | --- | --- | --- | --- |
|  |  | Bias | MBSE | ESE | CP | Bias | MBSE | ESE | CP |
| Outcome 1: |  |  |  |  |  |  |  |  |  |
| 5 | PALM | −0.055 | 0.384 | 0.356 | 0.932 | −0.057 | 0.382 | 0.368 | 0.937 |
|  | PALM-equal | 0.004 | 0.301 | 0.318 | 0.922 | −0.028 | 0.311 | 0.316 | 0.934 |
| 10 | PALM | −0.026 | 0.274 | 0.292 | 0.942 | −0.019 | 0.273 | 0.331 | 0.944 |
|  | PALM-equal | −0.001 | 0.201 | 0.232 | 0.905 | −0.021 | 0.207 | 0.224 | 0.921 |
| 15 | PALM | −0.008 | 0.222 | 0.218 | 0.939 | −0.006 | 0.221 | 0.236 | 0.933 |
|  | PALM-equal | 0.004 | 0.167 | 0.189 | 0.910 | −0.012 | 0.169 | 0.187 | 0.918 |
| 20 | PALM | −0.005 | 0.192 | 0.194 | 0.949 | 0.012 | 0.192 | 0.203 | 0.937 |
|  | PALM-equal | −0.004 | 0.145 | 0.157 | 0.936 | −0.012 | 0.147 | 0.160 | 0.935 |
| Outcome 2: |  |  |  |  |  |  |  |  |  |
| 5 | PALM | 0.024 | 0.377 | 0.336 | 0.941 | 0.037 | 0.376 | 0.329 | 0.953 |
|  | PALM-equal | −0.005 | 0.290 | 0.309 | 0.918 | 0.017 | 0.297 | 0.311 | 0.924 |
| 10 | PALM | 0.001 | 0.270 | 0.249 | 0.948 | 0.005 | 0.268 | 0.238 | 0.962 |
|  | PALM-equal | −0.002 | 0.205 | 0.231 | 0.907 | 0.003 | 0.206 | 0.226 | 0.920 |
| 15 | PALM | 0.002 | 0.221 | 0.204 | 0.955 | 0.000 | 0.219 | 0.199 | 0.958 |
|  | PALM-equal | −0.002 | 0.168 | 0.188 | 0.915 | 0.000 | 0.169 | 0.188 | 0.917 |
| 20 | PALM | −0.002 | 0.191 | 0.178 | 0.957 | −0.003 | 0.190 | 0.173 | 0.962 |
|  | PALM-equal | −0.002 | 0.146 | 0.156 | 0.939 | −0.002 | 0.147 | 0.162 | 0.928 |

**Table 2:** Comparison of the bias, model based standard error (MBSE), empirical standard error (ESE) and the coverage probability (CP) of the effect sizes of treatment comparison AB and AC estimated from the PALM and PALM-equal methods, under the Setting 2 where between-study variances were not equal.

| No. of studies | Method | Comparison AB |  |  |  | Comparison AC |  |  |  |
| --- | --- | --- | --- | --- | --- | --- | --- | --- | --- |
|  |  | Bias | MBSE | ESE | CP | Bias | MBSE | ESE | CP |
| Outcome 1: |  |  |  |  |  |  |  |  |  |
| 5 | PALM | −0.017 | 0.405 | 0.359 | 0.936 | −0.056 | 0.377 | 0.371 | 0.948 |
|  | PALM-equal | 0.008 | 0.288 | 0.332 | 0.889 | −0.028 | 0.305 | 0.331 | 0.908 |
| 10 | PALM | −0.060 | 0.288 | 0.257 | 0.945 | −0.038 | 0.286 | 0.272 | 0.951 |
|  | PALM-equal | 0.001 | 0.207 | 0.236 | 0.908 | −0.013 | 0.216 | 0.238 | 0.917 |
| 15 | PALM | −0.053 | 0.236 | 0.214 | 0.947 | −0.027 | 0.234 | 0.232 | 0.946 |
|  | PALM-equal | 0.002 | 0.236 | 0.190 | 0.918 | −0.008 | 0.234 | 0.197 | 0.917 |
| 20 | PALM | −0.055 | 0.204 | 0.189 | 0.946 | −0.026 | 0.203 | 0.204 | 0.948 |
|  | PALM-equal | 0.000 | 0.149 | 0.163 | 0.923 | −0.007 | 0.154 | 0.168 | 0.935 |
| Outcome 2: |  |  |  |  |  |  |  |  |  |
| 5 | PALM | 0.020 | 0.377 | 0.350 | 0.941 | 0.033 | 0.316 | 0.350 | 0.958 |
|  | PALM-equal | −0.003 | 0.300 | 0.333 | 0.901 | 0.012 | 0.290 | 0.310 | 0.924 |
| 10 | PALM | 0.013 | 0.269 | 0.256 | 0.941 | 0.014 | 0.266 | 0.228 | 0.964 |
|  | PALM-equal | 0.003 | 0.212 | 0.239 | 0.905 | 0.007 | 0.204 | 0.220 | 0.921 |
| 15 | PALM | 0.005 | 0.221 | 0.212 | 0.945 | 0.005 | 0.218 | 0.190 | 0.965 |
|  | PALM-equal | −0.001 | 0.221 | 0.196 | 0.914 | 0.002 | 0.218 | 0.181 | 0.927 |
| 20 | PALM | 0.005 | 0.192 | 0.185 | 0.945 | 0.002 | 0.189 | 0.165 | 0.966 |
|  | PALM-equal | 0.001 | 0.152 | 0.169 | 0.916 | 0.002 | 0.145 | 0.156 | 0.929 |

In summary, our proposed method assuming unequal between-study variances provides correct coverage probabilities under both settings, while the proposed algorithm assuming equal variance provides more accurate estimation in terms of bias, with slightly lower converge probabilities when sample size is small. Depending on the complexity of the network and number of studies for each contrast, treating *τ* ^2^ as equal can provide benefits in the accuracy and the computational efficiency of the estimation of effect sizes, while the converge of the confidence interval might be slightly smaller.

## 7 Discussion

In this study, we proposed a comprehensive framework, PALM, for multiple-outcome NMAs, including a robust and parsimonious modeling approach and a fast computational algorithm for estimation and inference. Our proposed method is advantageous over existing methods for large-scale multiple-outcome NMA (when the number of treatment and/or the number of outcomes is large) as it avoids the need to specify the complex between or within-study correlation structures. The iterative algorithm for parameter estimation is computationally efficient and it converges within seconds even for very large NMAs. We also proposed a comprehensive set of tools for comparing multiple treatments, including a new visualization tool named *star plot*, and an approach for patient-centered treatment ranking which facilitates evidence-based clinical decision making.

Our method is most beneficial when the population-level effect size ***µ*** is of interest, instead of the heterogeneity variance ***τ*** ^***2***^. Although we allow the heterogeneity variances to be different across treatment comparisons, estimating all these unique heterogeneity variances for different contrasts is challenging due to the often limited number of studies which directly compare specific treatments. This is a common problem in many NMA methods, and most of the current approaches assumed a common heterogeneity variance for all contrasts. However, the information orthogonality of our model formulation ensures that the accuracy of estimating ***µ*** is not sensitive to the assumptions regarding ***τ*** ^***2***^, and our simulation study demonstrates the benefits and validity of using the equal-variance model as a parsimonious working model.

The proposed method only requires specification the marginal distribution of each observation, which reduces the complexity of the model specification, and also improves the computational efficiency and the robustness in terms of misspecification of the correlation structures. Compared to existing Bayesian approaches which require specification of the correlation matrix and its prior (Efthimiou et al. 2014, 2015), one limitation of our method is that it does not allow borrowing information across outcomes or contrasts. When the correlation structure is known, or the within study correlations are reported in each study, our method is not able to incorporate such information, and therefore may lose some efficiency. However, as demonstrated in the setting of multivariate meta-analysis (Trikalinos et al. 2013), the efficiency gain from modeling correlation structures might be limited.

The potential violation of the consistency assumption, as well as the risk for publication bias and outcome reporting bias need to be evaluated to avoid misleading conclusions and inference. As mentioned in Section 3.4, we can modify the current model to allow evidence inconsistency, and by comparing with the model under consistency assumption, we may test and quantify the strength of inconsistency. Selection models and sensitivity analysis can be incorporated with the current model to correct the bias due to publication and outcome reporting bias (Copas and Shi 2000; Copas et al. 2014, 2019; Kirkham et al. 2012, 2018; Schmid 2017).

Recently, there have been a few important efforts in collecting published networks and creating databases for NMAs: Nikolakopoulou et al. (2014) created a database containing 186 networks of interventions, and Petropoulou et al. (2017) collected data for 456 NMAs published between 1999 to 2015. These databases provide opportunities for evaluating our method across large number of NMAs to compare with existing methods, and updating results for patient-centered treatment ranking. Following their efforts, we are collecting NMAs published after 2015, and till the submission of this manuscript, we have collected additional 152 NMA studies where a large portion of these studies reported more than two outcomes. We are evaluating the broad impact of our method on a large number of NMA networks, and the results will be summarized and reported in the near future.

## Data Availability

Our analysis is based on public available dataset from published papers of randomized clinical trials.

https://doi.org/10.1136/bmj.h217

## Footnotes

† See Appendix G of the Supplementary Material.

## References

Alfirevic, Z., E. Keeney, T. Dowswell, N. J. Welton, S. Dias, L. V. Jones, K. Navaratnam, and D. M. Caldwell (2015). Labour induction with prostaglandins: a systematic review and network meta-analysis. BMJ 350, h217.

Bahji, A., D. Ermacora, C. Stephenson, E. R. Hawken, and G. Vazquez (2020). Comparative efficacy and tolerability of pharmacological treatments for the treatment of acute bipolar depression: a systematic review and network meta-analysis. Journal of Affective Disorders.

Bash, L. D., J. L. Buono, G. M. Davies, A. Martin, K. Fahrbach, H. Phatak, R. Avetisyan, and M. Mwamburi (2012). Systematic review and meta-analysis of the efficacy of cardioversion by vernakalant and comparators in patients with atrial fibrillation. Cardiovascular drugs and therapy 26 (2), 167–179.

Boulvain, M., A. J. Kelly, and O. Irion (2008). Intracervical prostaglandins for induction of labour. Cochrane Database of Systematic Reviews 1, Art. No.: CD006971.

Bygdeman, M. (2003). Pharmacokinetics of prostaglandins. Best Practice & Research Clinical Obstetrics & Gynaecology 17 (5), 707–716.

Caldwell, D. M., A. E. Ades, and J. P. T. Higgins (2005). Simultaneous comparison of multiple treatments: combining direct and indirect evidence. BMJ 331 (7521), 897–900.

Caughey, A. B., A. G. Cahill, J.-M. Guise, and D. J. Rouse (2014). Safe prevention of the primary cesarean delivery. American Journal of Obstetrics and Gynecology 210 (3), 179–193.

Centers for Disease Control and Prevention (2020 (accessed February 3, 2020)). National Center for Health Statistics–Births.

Chaimani, A., J. P. T. Higgins, D. Mavridis, P. Spyridonos, and G. Salanti (2013). Graphical tools for network meta-analysis in stata. PLOS ONE 8 (10), e76654.

Cipriani, A., T. A. Furukawa, G. Salanti, A. Chaimani, L. Z. Atkinson, Y. Ogawa, S. Leucht, H. G. Ruhe, E. H. Turner, J. P. T. Higgins, M. Egger, N. Takeshima, Y. Hayasaka, H. Imai, K. Shinohara, A. Tajika, J. P. A. Ioannidis, and J. R. Geddes (2018). Comparative efficacy and acceptability of 21 antidepressant drugs for the acute treatment of adults with major depressive disorder: a systematic review and network meta-analysis. The Lancet 391 (10128), 1357–1366.

Copas, J., K. Dwan, J. Kirkham, and P. Williamson (2014). A model-based correction for outcome reporting bias in meta-analysis. Biostatistics 15 (2), 370–383.

Copas, J., A. Marson, P. Williamson, and J. Kirkham (2019). Model-based sensitivity analysis for outcome reporting bias in the meta analysis of benefit and harm outcomes. Statistical methods in medical research 28 (3), 889–903.

Copas, J. and J. Q. Shi (2000). Meta-analysis, funnel plots and sensitivity analysis. Biostatistics 1 (3), 247–262.

Cox, D. R. and N. Reid (2004). A note on pseudolikelihood constructed from marginal densities. Biometrika 91 (3), 729–737.

Davey, J., R. M. Turner, M. J. Clarke, and J. P. T. Higgins (2011). Characteristics of meta-analyses and their component studies in the Cochrane Database of Systematic Reviews: a cross-sectional, descriptive analysis. BMC Medical Research Methodology 11 (1), 160.

Dias, S., N. J. Welton, A. J. Sutton, D. M. Caldwell, G. Lu, and A. E. Ades (2013). Evidence synthesis for decision making 4: inconsistency in networks of evidence based on randomized controlled trials. Medical Decision Making 33 (5), 641–656.

Dulai, P. S., S. Singh, E. Marquez, R. Khera, L. J. Prokop, P. J. Limburg, S. Gupta, and M. H. Murad (2016). Chemoprevention of colorectal cancer in individuals with previous colorectal neoplasia: systematic review and network meta-analysis. Bmj 355, i6188.

Dunkley, A., K. Charles, L. Gray, J. Camosso-Stefinovic, M. Davies, and K. Khunti (2012). Effectiveness of interventions for reducing diabetes and cardiovascular disease risk in people with metabolic syndrome: systematic review and mixed treatment comparison meta-analysis. Diabetes, Obesity and Metabolism 14 (7), 616–625.

Efthimiou, O., D. Mavridis, A. Cipriani, S. Leucht, P. Bagos, and G. Salanti (2014). An approach for modelling multiple correlated outcomes in a network of interventions using odds ratios. Statistics in Medicine 33 (13), 2275–2287.

Efthimiou, O., D. Mavridis, R. D. Riley, A. Cipriani, and G. Salanti (2015). Joint synthesis of multiple correlated outcomes in networks of interventions. Biostatistics 16 (1), 84–97.

Guyatt, G. H., A. D. Oxman, R. Kunz, J. Woodcock, J. Brozek, M. Helfand, P. Alonso-Coello, Y. Falck-Ytter, R. Jaeschke, G. Vist, E. A. Akl, P. N. Post, S. Norris, J. Meerpohl, V. K. Shukla, M. Nasser, and H. J. SchuÜnemann (2011). GRADE guidelines: 8. rating the quality of evidence—indirectness. Journal of Clinical Epidemiology 64 (12), 1303–1310.

Jackson, D., S. Bujkiewicz, M. Law, R. D. Riley, and I. R. White (2018). A matrix-based method of moments for fitting multivariate network meta-analysis models with multiple outcomes and random inconsistency effects. Biometrics 74 (2), 548–556.

Jackson, D., R. Riley, and I. R. White (2011). Multivariate meta-analysis: potential and promise. Statistics in Medicine 30 (20), 2481–2498.

Khan, R.-U., H. El-Refaey, S. Sharma, D. Sooranna, and M. Stafford (2004). Oral, rectal, and vaginal pharmacokinetics of misoprostol. Obstetrics & Gynecology 103 (5), 866–870.

Kirkham, J. J., D. G. Altman, A.-W. Chan, C. Gamble, K. M. Dwan, and P. R. Williamson (2018). Outcome reporting bias in trials: a methodological approach for assessment and adjustment in systematic reviews. bmj 362, k3802.

Kirkham, J. J., R. D. Riley, and P. R. Williamson (2012). A multivariate meta-analysis approach for reducing the impact of outcome reporting bias in systematic reviews. Statistics in medicine 31 (20), 2179–2195.

Li, F., W. Huang, and X. Zhang (2018). Efficacy and safety of different regimens for primary open-angle glaucoma or ocular hypertension: a systematic review and network meta-analysis. Acta ophthalmologica 96 (3), e277–e284.

Liang, K.-Y. (1983). On information and ancillarity in the presence of a nuisance parameter. Biometrika 70 (3), 607–612.

Liang, K.-Y., S. L. Zeger, et al. (1995). Inference based on estimating functions in the presence of nuisance parameters. Statistical Science 10 (2), 158–173.

Lindsay, B. G. (1988). Composite likelihood methods. Contemporary Mathematics 80 (1), 221– 239.

Liu, Y., S. M. DeSantis, and Y. Chen (2018). Bayesian mixed treatment comparisons meta-analysis for correlated outcomes subject to reporting bias. Journal of the Royal Statistical Society: Series C (Applied Statistics) 67 (1), 127–144.

Lu, G. and A. E. Ades (2006). Assessing evidence inconsistency in mixed treatment comparisons. Journal of the American Statistical Association 101 (474), 447–459.

Lu, G. and A. E. Ades (2009). Modeling between-trial variance structure in mixed treatment comparisons. Biostatistics 10 (4), 792–805.

Lumley, T. (2002). Network meta-analysis for indirect treatment comparisons. Statistics in Medicine 21 (16), 2313–2324.

Mauri, D., N. P. Polyzos, G. Salanti, N. Pavlidis, and J. P. Ioannidis (2008). Multiple-treatments meta-analysis of chemotherapy and targeted therapies in advanced breast cancer. JNCI: Journal of the National Cancer Institute 100 (24), 1780–1791.

Mavridis, D., R. Porcher, A. Nikolakopoulou, G. Salanti, and P. Ravaud (2019). Extensions of the probabilistic ranking metrics of competing treatments in network meta-analysis to reflect clinically important relative differences on many outcomes. Biometrical Journal.

Nikolakopoulou, A., A. Chaimani, A. A. Veroniki, H. S. Vasiliadis, C. H. Schmid, and G. Salanti (2014). Characteristics of networks of interventions: a description of a database of 186 published networks. PLOS ONE 9 (1), e86754.

Petropoulou, M., A. Nikolakopoulou, A.-A. Veroniki, P. Rios, A. Vafaei, W. Zarin, M. Giannatsi, S. Sullivan, A. C. Tricco, A. Chaimani, et al. (2017). Bibliographic study showed improving statistical methodology of network meta-analyses published between 1999 and 2015. Journal of clinical epidemiology 82, 20–28.

Pillinger, T., R. A. McCutcheon, L. Vano, Y. Mizuno, A. Arumuham, G. Hindley, K. Beck, S. Nate- san, O. Efthimiou, A. Cipriani, et al. (2020). Comparative effects of 18 antipsychotics on metabolic function in patients with schizophrenia, predictors of metabolic dysregulation, and association with psychopathology: a systematic review and network meta-analysis. The Lancet Psychiatry 7 (1), 64–77.

Riley, R. D., D. Jackson, G. Salanti, D. L. Burke, M. Price, J. Kirkham, and I. R. White (2017). Multivariate and network meta-analysis of multiple outcomes and multiple treatments: rationale, concepts, and examples. BMJ 358, j3932.

Riley, R. D., J. R. Thompson, and K. R. Abrams (2007). An alternative model for bivariate random-effects meta-analysis when the within-study correlations are unknown. Biostatistics 9 (1), 172– 186.

Salanti, G. (2012). Indirect and mixed-treatment comparison, network, or multiple-treatments meta-analysis: many names, many benefits, many concerns for the next generation evidence synthesis tool. Research Synthesis Methods 3 (2), 80–97.

Salanti, G., A. Ades, and J. P. Ioannidis (2011). Graphical methods and numerical summaries for presenting results from multiple-treatment meta-analysis: an overview and tutorial. Journal of clinical epidemiology 64 (2), 163–171.

Salanti, G., J. P. T. Higgins, A. E. Ades, and J. P. A. Ioannidis (2008). Evaluation of networks of randomized trials. Statistical Methods in Medical Research 17 (3), 279–301.

Schmid, C. H. (2017). Outcome reporting bias: a pervasive problem in published meta-analyses. American Journal of Kidney Diseases 69 (2), 172–174.

Silver, R. M. (2012). Implications of the first cesarean: perinatal and future reproductive health and subsequent cesareans, placentation issues, uterine rupture risk, morbidity, and mortality. Seminars in Perinatology 36 (5), 315–323.

Slee, A., I. Nazareth, P. Bondaronek, Y. Liu, Z. Cheng, and N. Freemantle (2019). Pharmacological treatments for generalised anxiety disorder: a systematic review and network meta-analysis. The Lancet 393 (10173), 768–777.

Tang, O. S., H. Schweer, H. Seyberth, S. W. Lee, and P. C. Ho (2002). Pharmacokinetics of different routes of administration of misoprostol. Human Reproduction 17 (2), 332–336.

Terasawa, T., N. A. Trikalinos, B. Djulbegovic, and T. A. Trikalinos (2013). Comparative efficacy of first-line therapies for advanced-stage chronic lymphocytic leukemia: a multiple-treatment meta-analysis. Cancer treatment reviews 39 (4), 340–349.

Trikalinos, T. A., D. C. Hoaglin, and C. H. Schmid (2013). Empirical and simulation-based comparison of univariate and multivariate meta-analysis for binary outcomes.

Trinquart, L., N. Attiche, A. Bafeta, R. Porcher, and P. Ravaud (2016). Uncertainty in treatment rankings: reanalysis of network meta-analyses of randomized trials. Annals of internal medicine 164 (10), 666–673.

White, I. R., J. K. Barrett, D. Jackson, and J. P. T. Higgins (2012). Consistency and inconsistency in network meta-analysis: model estimation using multivariate meta-regression. Research Synthesis Methods 3 (2), 111–125.

Zhu, B., N. Song, R. Shen, A. Arora, M. J. Machiela, L. Song, M. T. Landi, D. Ghosh, N. Chatterjee, V. Baladandayuthapani, et al. (2017). Integrating clinical and multiple omics data for prognostic assessment across human cancers. Scientific reports 7 (1), 1–13.

