## Supplementary Material for "PALM: Patient-centered Treatment Ranking via Large-scale Multivariate Network Meta-analysis"

This supplementary file includes the detailed form of the variance estimator of the proposed method (Appendix A); the estimation algorithm and the variance estimator of the model assuming equal between-study variances across treatment comparisons (Appendix B); the modified formulation of the model to relax the consistency assumption (Appendix C); an example to show the limitation of the radar plot (Appendix D), which motivated our proposed star plot; additional results for the labor induction NMA (Appendix E); and regularity conditions and the sketch proof of Theorem 1 (Appendix F).

#### Appendix A: Variance Estimation

We define

$$\hat{I} = -\frac{\partial^2 L(\eta)}{\partial \eta^2}, \tag{1}$$

and denote

$$S_i^{XY} = \partial \Big\{ \log\left( s_{k,i}^{\mathrm{XY}2} + \tau_k^{\mathrm{XY}2} \right) + \frac{\left( y_{k,i}^{\mathrm{XY}} - \mu_k^{\mathrm{XY}} \right)^2}{s_{k,i}^{\mathrm{XY}2} + \tau_k^{\mathrm{XY}2}} \Big\} / \partial \eta.$$

Define

$$\hat{\Lambda} = \frac{1}{4} \sum_{k=1}^{2} \left\{ \sum_{\mathrm{XY} \in \{\mathrm{AB},\mathrm{BC},\mathrm{AC}\}} \sum_{i \in \mathcal{N}_{\mathrm{XY}}^k} S_i^{XY} S_i^{XY\,\mathrm{T}} \right\}. \tag{2}$$

The variance of the estimator $\hat{\eta}$ from Equation (5) of the main paper can be estimated by the following estimator

$$\hat{V}(\hat{\eta}) = \hat{I}^{-1} \hat{\Lambda} \hat{I}^{-1}. \tag{3}$$

#### Appendix B: Model Assuming Equal Between-Study Variances

When assuming equal between-study variance for each outcome, i.e., $\tau_k^{\mathrm{AB}} = \tau_k^{\mathrm{AC}} = \tau_k^{\mathrm{BC}}$ for $k = 1, 2$, Algorithm 1 can be slightly modified. First, we define the log-pseudolikelihood function for each outcome,

$$L_k(\boldsymbol{\mu}_k, \tau_k^2) = \sum_{\text{XY}\in\{\text{AB,BC,AC}\}} \sum_{i\in\mathcal{N}_{\text{XY}}^k} \left[ \log\left( s_{k,i}^{\text{XY}^2} + \tau_k^2 \right) + \frac{\left( y_{k,i}^{\text{XY}} - \mu_k^{\text{XY}} \right)^2}{s_{k,i}^{\text{XY}^2} + \tau_k^2} \right].$$

And for a given $\tau_k^{2(t)}$, we denote

$$w_{k,i}^{\text{XY}(t)} = \left( s_{k,i}^{\text{XY}^2} + \tau_k^{2(t)} \right)^{-1};$$

$$\mathbf{H}_k^{(t)} = \begin{pmatrix} \sum_{i\in\mathcal{N}_{\text{AB}}^k} w_{k,i}^{\text{AB}(t)} + \sum_{i\in\mathcal{N}_{\text{BC}}^k} w_{k,i}^{\text{BC}(t)} & -\sum_{i\in\mathcal{N}_{\text{BC}}^k} w_{k,i}^{\text{BC}(t)} \\ -\sum_{i\in\mathcal{N}_{\text{BC}}} w_{k,i}^{\text{BC}(t)} & \sum_{i\in\mathcal{N}_{\text{AC}}^k} w_{k,i}^{\text{AC}(t)} + \sum_{i\in\mathcal{N}_{\text{BC}}^k} w_{k,i}^{\text{BC}(t)} \end{pmatrix};$$

$$\boldsymbol{v}_k^{(t)} = \begin{pmatrix} \sum_{i\in\mathcal{N}_{\text{AB}}^k} y_{k,i}^{\text{AB}} w_{k,i}^{\text{AB}(t)} - \sum_{i\in\mathcal{N}_{\text{BC}}^k} y_{k,i}^{\text{BC}} w_{k,i}^{\text{BC}(t)} \\ \sum_{i\in\mathcal{N}_{\text{AC}}^k} y_{k,i}^{\text{AC}} w_{k,i}^{\text{AC}(t)} + \sum_{i\in\mathcal{N}_{\text{BC}}^k} y_{k,i}^{\text{BC}} w_{k,i}^{\text{BC}(t)} \end{pmatrix}.$$

Maximizing $L_k\left(\boldsymbol{\mu}_k, \tau_k^{2(t)}\right)$ over $\boldsymbol{\mu}_k$ yields

$$\mathbf{H}_k^{(t)} \boldsymbol{\mu}_k = \boldsymbol{v}_k^{(t)}.$$

Thus, Algorithm 1 can be changed to the following algorithm.

**Algorithm 1** Iterative estimation

**for** $k = 1, 2$ **do**
  Specify the initial values $\boldsymbol{\tau}_k^{(0)}$, $D = 1$.
  **while** $t \leq T$ or $D > \delta$, **do**
    Obtain $\boldsymbol{\mu}_k^{(t+1)}$ by
$$\boldsymbol{\mu}_k^{(t+1)} = \left(\mathbf{H}_k^{(t)}\right)^{-1} \boldsymbol{v}_k^{(t)}$$
    Obtain $\boldsymbol{\tau}_k^{(t+1)}$ by
$$\boldsymbol{\tau}_k^{(t+1)} = \arg\max_{\boldsymbol{\tau}_k} L_k\left(\boldsymbol{\mu}_k^{(t+1)}, \boldsymbol{\tau}_k^2\right)$$
    Update $D = \left|\boldsymbol{\tau}_k^{(t+1)} - \boldsymbol{\tau}_k^{(t)}\right|$ and $t = t + 1$
  **end**
**end**
**return** $\boldsymbol{\mu}_1^{(T)}, \boldsymbol{\tau}_1^{(T)}, \boldsymbol{\mu}_2^{(T)}, \boldsymbol{\tau}_2^{(T)}$.

To estimate the variance of the estimator from the above algorithm, we define $\boldsymbol{\eta} = (\boldsymbol{\mu}^{\mathrm{T}}, \tau_1^2, \tau_2^2)^{\mathrm{T}}$, and define

$$L_{\text{equal}} = L_1(\boldsymbol{\mu}_1, \tau_1^2) + L_2(\boldsymbol{\mu}_1, \tau_2^2).$$

$$\tilde{I} = -\frac{\partial^2 L_{\text{equal}}(\boldsymbol{\eta})}{\partial \boldsymbol{\eta}^2}, \tag{4}$$

and denote

$$S_i^{XY} = \partial \Big\{ \log \left( s_{k,i}^{\mathrm{XY}\,2} + {\tau_k}^2 \right) + \frac{\left(y_{k,i}^{\mathrm{XY}} - \mu_k^{\mathrm{XY}}\right)^2}{s_{k,i}^{\mathrm{XY}\,2} + {\tau_k}^2} \Big\} / \partial \eta.$$

Define

$$\tilde{\Lambda} = \frac{1}{4} \sum_{k=1}^{2} \left\{ \sum_{\mathrm{XY} \in \{\mathrm{AB,BC,AC}\}} \sum_{i \in \mathcal{N}_{\mathrm{XY}}^k} S_i^{XY} S_i^{XY\,\mathrm{T}} \right\}. \tag{5}$$

The variance of the estimator $\tilde{\eta}$ obtained by maximizing $L_{\text{equal}}$ can be estimated by the following estimator

$$\tilde{V}(\tilde{\eta}) = \tilde{I}^{-1} \tilde{\Lambda} \tilde{I}^{-1}. \tag{6}$$

#### Appendix C: Inconsistency Formulation

When the consistency assumption does not hold, i.e., we are not confident to assume that

$$\mu_k^{\mathrm{BC}} = \mu_k^{\mathrm{AC}} - \mu_k^{\mathrm{AB}} \text{ for } k = 1, 2.$$

We will treat $\mu_k^{\mathrm{BC}}$, $\mu_k^{\mathrm{AC}}$ and $\mu_k^{\mathrm{AB}}$ as free parameters. Denote $\boldsymbol{\mu} = (\mu_1^{\mathrm{AB}}, \mu_1^{\mathrm{BC}}, \mu_1^{\mathrm{AC}}, \mu_2^{\mathrm{AB}}, \mu_2^{\mathrm{BC}}, \mu_2^{\mathrm{AC}})^{\mathrm{T}}$, and $\boldsymbol{\eta} = (\boldsymbol{\mu}^{\mathrm{T}}, \boldsymbol{\tau}^{2\mathrm{T}})^{\mathrm{T}}$, we can obtain the estimator of $\boldsymbol{\eta}$ by optimizing the following log composite likelihood function:

$$L(\boldsymbol{\eta}) = -\frac{1}{2} \sum_{k=1}^{2} \left\{ \sum_{\mathrm{XY} \in \{\mathrm{AB,BC,AC}\}} \sum_{i \in \mathcal{N}_{\mathrm{XY}}^k} \left[ \log \left( s_{k,i}^{\mathrm{XY}\,2} + \tau_k^{\mathrm{XY}\,2} \right) + \frac{\left(y_{k,i}^{\mathrm{XY}} - \mu_k^{\mathrm{XY}}\right)^2}{s_{k,i}^{\mathrm{XY}\,2} + \tau_k^{\mathrm{XY}\,2}} \right] \right\}. \tag{7}$$

Since now each comparison has their own set of parameters, optimizing the above function is equivalent as optimizing separately the following functions

$$L_k^{\mathrm{XY}}(\mu_k^{\mathrm{XY}}, \tau_k^{\mathrm{XY}2}) = -\frac{1}{2}\sum_{i\in\mathcal{N}_{\mathrm{XY}}^k}\left[\log\left(s_{k,i}^{\mathrm{XY}2} + \tau_k^{\mathrm{XY}2}\right) + \frac{\left(y_{k,i}^{\mathrm{XY}} - \mu_k^{\mathrm{XY}}\right)^2}{s_{k,i}^{\mathrm{XY}2} + \tau_k^{\mathrm{XY}2}}\right] \quad (8)$$

$for \mathrm{XY} \in \{\mathrm{AB}, \mathrm{BC}, \mathrm{AC}\}$, and $k \in 1, 2$, which is equivalent as fitting a random-effects model for each treatment comparison and each outcome. However, now without the consistency assumption, only the effect sizes of comparisons being compared by randomized clinical trials can be estimated by the above procedure.

When assuming equal variance, the pseudo likelihood function can be written as

$$L_{\text{equal}}(\boldsymbol{\eta}) = -\frac{1}{2}\sum_{k=1}^{2}\left\{\sum_{\mathrm{XY}\in\{\mathrm{AB,BC,AC}\}}\sum_{i\in\mathcal{N}_{\mathrm{XY}}^k}\left[\log\left(s_{k,i}^{\mathrm{XY}2} + {\tau_k}^2\right) + \frac{\left(y_{k,i}^{\mathrm{XY}} - \mu_k^{\mathrm{XY}}\right)^2}{s_{k,i}^{\mathrm{XY}2} + {\tau_k}^2}\right]\right\}. \quad (9)$$

which can be decomposed to

$$L_{\text{equal}}(\boldsymbol{\eta}) = -\frac{1}{2}\sum_{k=1}^{2} L_k(\boldsymbol{\mu}_k, \tau_k^2) \quad (10)$$

where

$$L_k(\boldsymbol{\mu}_k, \tau_k^2) = \sum_{\mathrm{XY}\in\{\mathrm{AB,BC,AC}\}}\sum_{i\in\mathcal{N}_{\mathrm{XY}}^k}\left[\log\left(s_{k,i}^{\mathrm{XY}2} + {\tau_k}^2\right) + \frac{\left(y_{k,i}^{\mathrm{XY}} - \mu_k^{\mathrm{XY}}\right)^2}{s_{k,i}^{\mathrm{XY}2} + {\tau_k}^2}\right] \quad (11)$$

We observe that for a given $\tau_k^{2(t)}$, $\mu_k^{\mathrm{XY}}$ can be calculated as

$$\mu_k^{\mathrm{XY}(t+1)} = \frac{\sum_{i\in\mathcal{N}_{\mathrm{AB}}^k} y_{k,i}^{\mathrm{XY}} w_{k,i}^{\mathrm{AB}(t)}}{\sum_{i\in\mathcal{N}_{\mathrm{AB}}^k} w_{k,i}^{\mathrm{AB}(t)}} \quad (12)$$

where $w_{k,i}^{\mathrm{XY}(t)} = \left(s_{k,i}^{\mathrm{XY}2} + \tau_k^{2(t)}\right)^{-1}$; We have the following algorithm

#### Appendix D: Limitation of the Radar Plot

The radar chart, which is commonly used for comparing multiple outcomes (Zhu et al. 2017), has a limitation in the setting of treatment comparison. As shown in Figure S1, we have two treatments (A and B) and we want to compare them over five outcomes using a radar plot. In the upper panel, five outcomes from outcome1 to outcome 5 are ordered in clockwise order. The area of shaded region of treatment A is apparently larger than that of treatment B. However, if we swap the two

**Algorithm 2** Iterative estimation for inconsistency model with equal heterogeneity variance

**for** $k = 1, 2$ **do**
  Specify the initial values $\boldsymbol{\tau}_k^{(0)}$, $D = 1$.
  **while** $t \leq T$ or $D > \delta$, **do**
    Obtain $\boldsymbol{\mu}_k^{(t+1)}$ by

$$\mu_k^{\text{XY}(t+1)} = \frac{\sum_{i \in \mathcal{N}_{\text{AB}}^k} y_{k,i}^{\text{XY}} w_{k,i}^{\text{AB}(t)}}{\sum_{i \in \mathcal{N}_{\text{AB}}^k} w_{k,i}^{\text{AB}(t)}}$$

    Obtain $\boldsymbol{\tau}_k^{(t+1)}$ by

$$\boldsymbol{\tau}_k^{(t+1)} = \arg\max_{\boldsymbol{\tau}_k} L_k \left( \boldsymbol{\mu}_k^{(t+1)}, \boldsymbol{\tau}_k^2 \right)$$

    Update $D = \left| \boldsymbol{\tau}_k^{(t+1)} - \boldsymbol{\tau}_k^{(t)} \right|$ and $t = t + 1$
  **end**
**end**
**return** $\boldsymbol{\mu}_1^{(T)}, \boldsymbol{\tau}_1^{(T)}, \boldsymbol{\mu}_2^{(T)}, \boldsymbol{\tau}_2^{(T)}$.

axes for outcome 4 and outcome 5 (lower panel), then the area of shaded region of treatment A becomes smaller than that of treatment B. Therefore, the area of the shaded region is sensitive to how the axes corresponding to the outcomes are ordered, which is the motivation of the star plot.

#### Appendix E: Additional Results for Labor Induction NMA

##### Model fitting results:

In this subsection, we show the complete model fitting results of our proposed method applied on the labor induction NMA. Figure S2 shows the estimated odds ratios with their 95% confidence intervals of the 13 treatments compared to placebo across the five outcomes. Figures S3-S7 show the head-to-head comparison of the 13 treatments the five outcomes.

##### WSUCRA based on a utility function

Please see Table S1 for the estimated WSCURA for the eight treatments corresponding to the three utility functions defined in Section 5 of the main paper. We also considered another utility function, which was constructed by a weighted sum of all 5 outcomes (See Figure S8).

**Weighted star plot**

Figure S9 shows the weighted star plot to compare 8 treatment options for labor induction across five outcomes, where outcomes hyperstimulation and vaginal delivery not within 24 hours were given half of the weight as the rest three outcomes. Based on this weighted star plot, we know that low dose of oral misoprostol solution was the best treatment when consider hyperstimulation and vaginal delivery not within 24 hours less important as cesarean section, maternal morbidity and neonatal morbidity.

**Sensitivity analysis**

In the sensitivity analysis, we only included the 162 studies out of the 280 studies which were considered in Alfirevic et al. (2015) as of low risk for bias. We show the forest plot of the estimating results in Figure S10.

#### Appendix F: Regularity Conditions and Proof of Theorem 1

**Regularity Conditions**

(C1). The parameter space, denoted by $\mathbf{\Theta}$ is compact and true parameter value, denoted by $\boldsymbol{\eta}^*$ is an interior point of $\mathbf{\Theta}$.

(C2). As $n \to \infty$, we assume the proportion of studies that report the estimated effect size and standard error of treatment comparison $XY$ for outcome $k$, i.e., $n_{\mathrm{XY}}^k$ satisfies $n_{\mathrm{XY}}^k/n \to r_{\mathrm{XY}}^k > 0$.

(C3). The marginal population risk, defined as

$$l_k^{XY}(\mu_k, \tau_k^2) = -\mathbb{E} \sum_{i \in \mathcal{N}_{\mathrm{XY}}^k} \left\{ \log\left( s_{k,i}^{\mathrm{XY}^2} + \tau_k^2 \right) + \frac{\left( y_{k,i}^{\mathrm{XY}} - \mu_k^{\mathrm{XY}} \right)^2}{s_{k,i}^{\mathrm{XY}^2} + \tau_k^2} \right\},$$

has an unique maximizer, which is the true parameter value $\{\mu_k^*, {\tau_k^*}^2\}$.

(C4). The first, second, and third order derivatives of $L(\boldsymbol{\eta})$ defined equation (4) of the main paper are bounded.

(C5). The expectation of the second-order derivation of $L(\boldsymbol{\eta})$ , i.e., $\mathbb{E}\nabla^2 L(\boldsymbol{\eta})$ exist and is positive definite.

**Sketch proof of Theorem 1** For each study, we define

$$h_i(\boldsymbol{\eta}) = -\sum_{k=1}^{2} \sum_{\text{XY}\in\{\text{AB,BC,AC}\}} \left\{ \log\left(s_{k,i}^{\text{XY}2} + \tau_k^2\right) + \frac{\left(y_{k,i}^{\text{XY}} - \mu_k^{\text{XY}}\right)^2}{s_{k,i}^{\text{XY}2} + \tau_k^2} \right\} I_{ik,\text{XY}}$$

where $I_{ik,\text{XY}}$ is an indicator of whether the $k$-th outcome of the $XY$ comparison is reported in the $i$-th study. According to C(3), the first derivative of $h_i(\boldsymbol{\eta})$, denoted by $\nabla h_i(\boldsymbol{\eta})$ is an unbiased estimating equation of $\boldsymbol{\eta}$. We noticed that $L(\boldsymbol{\eta})$ can be written as

$$L(\boldsymbol{\eta}) = \sum_{i=1}^{n} h_i(\boldsymbol{\eta}).$$

Thus, the maximum pseudolikelihood estimator of $L(\boldsymbol{\eta})$ is also the solution of the following estimating equation

$$\nabla L(\boldsymbol{\eta}) = \sum_{i=1}^{n} \nabla h_i(\boldsymbol{\eta}) = 0.$$

As the data in each study are independently distributed. using assumption (C1) - (C5), and the classical theory of unbiased estimating equations (e.g. Song and Song (2007)), we can show the asymptotic normality of the proposed estimator follows Theorem 1.

#### Appendix G: Calculation of Unknown Parameters

Here we give a detailed calculation of how we determine the number of unknown parameters in the between-study variance-covariance matrix mentioned in Section 2 of the main paper.

Since we have 5 outcomes for 13 treatment comparisons (setting placebo as the reference drug), there are in total $5 = 65$ different effect sizes. The unstructured variance-covariance matrix will have $\binom{65}{2} + 65 = 2145$ unique parameters since it is symmetric. Under a consistency assumption, the correlation between treatment comparisons for a specific outcome is often assumed known or can be derived from other parameters, so the number of unknown parameters can be reduced to $2145 - \binom{13}{2} \times 5 = 1755$.

### References

Alfirevic, Z., E. Keeney, T. Dowswell, N. J. Welton, S. Dias, L. V. Jones, K. Navaratnam, and D. M. Caldwell (2015). Labour induction with prostaglandins: a systematic review and network meta-analysis. *BMJ 350*, h217.

Song, X.-K. and P. X.-K. Song (2007). *Correlated data analysis: modeling, analytics, and applications*. Springer Science & Business Media.

Zhu, B., N. Song, R. Shen, A. Arora, M. J. Machiela, L. Song, M. T. Landi, D. Ghosh, N. Chatterjee, V. Baladandayuthapani, et al. (2017). Integrating clinical and multiple omics data for prognostic assessment across human cancers. *Scientific reports 7*(1), 1–13.

Figure S1: Example of changing the order of outcomes in the radar chart can lead to inconsistent ranking of treatment.

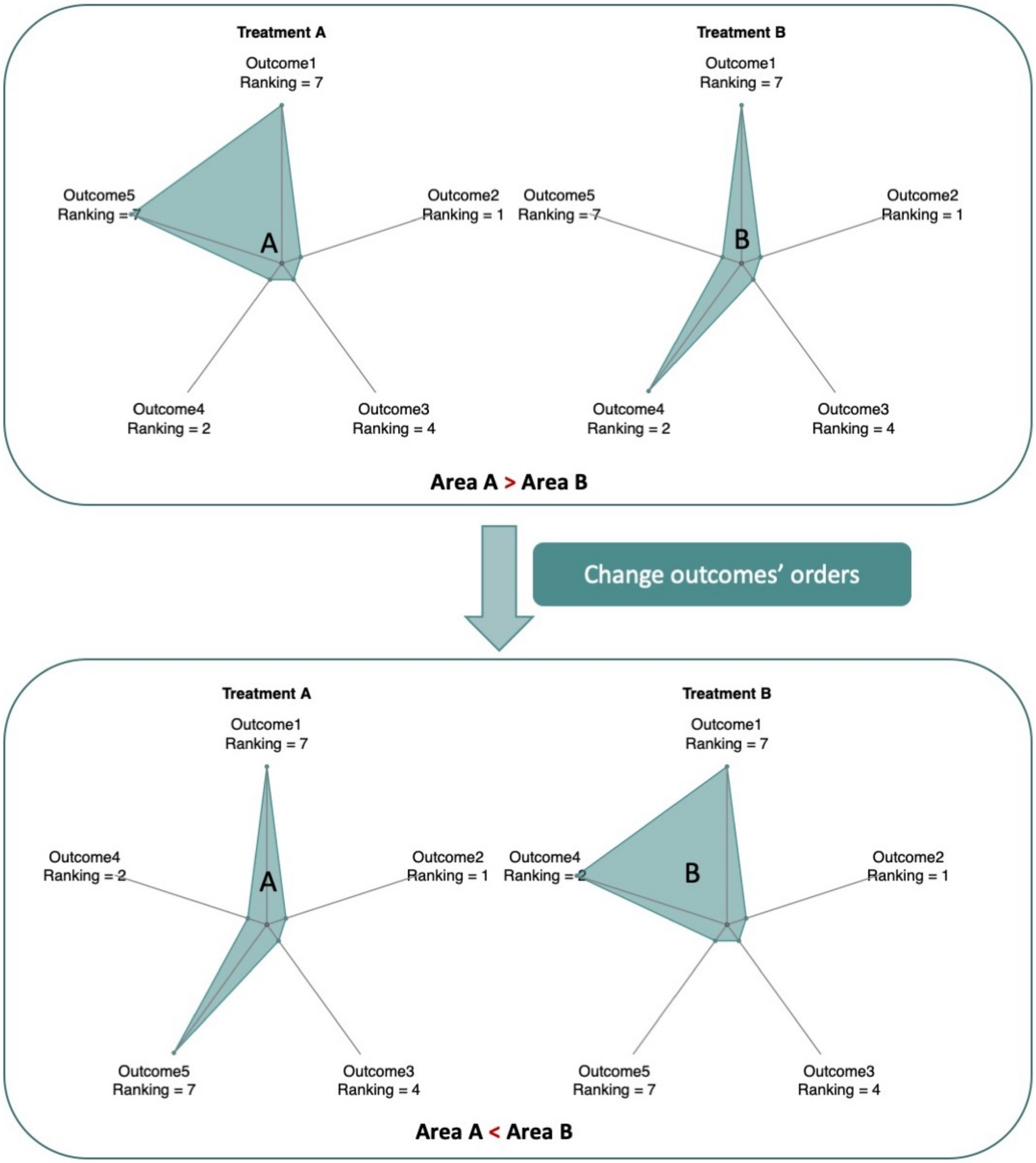

Figure S2: Forest plots of estimated odds ratios of the 13 labor induction treatment options across five outcomes.

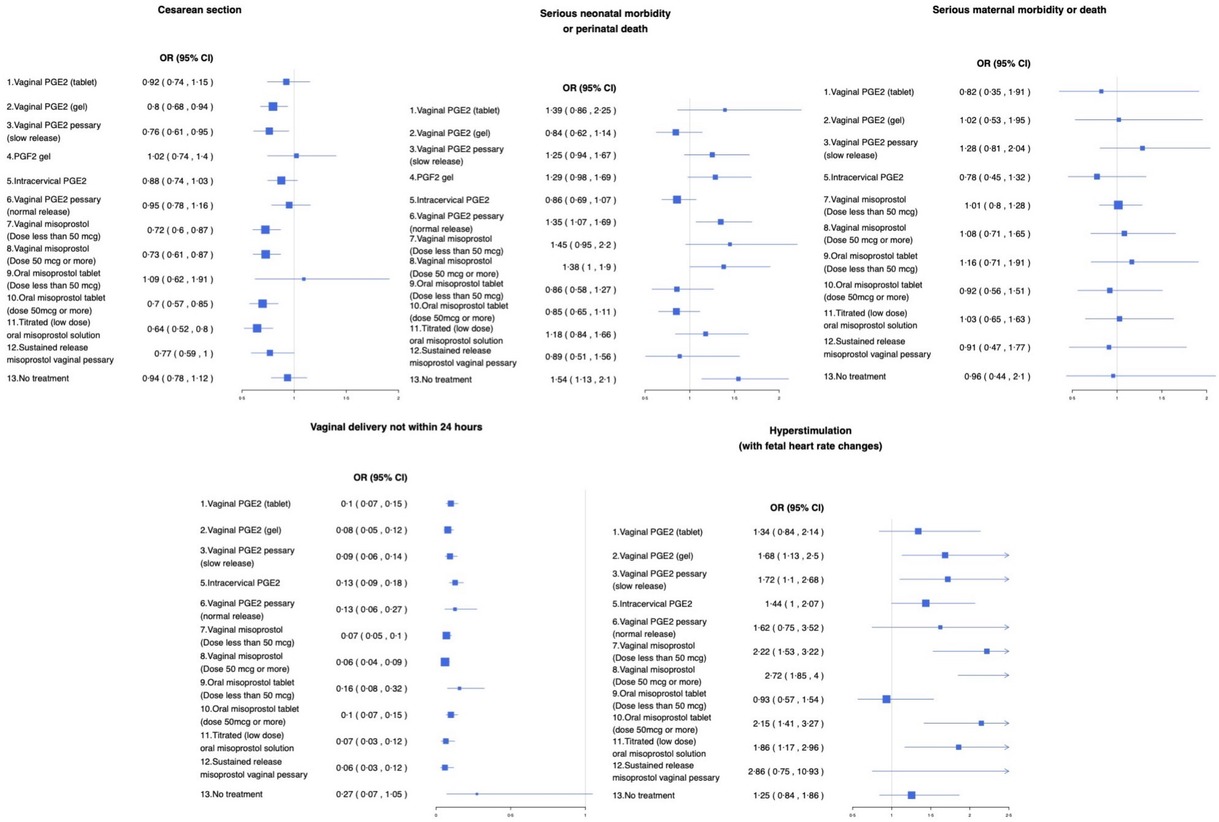

Figure S3: Forest plots of estimated odds ratios of the 13 labor induction treatment options across five outcomes.

**Outcome1:**
Cesarean section

| | | | | | | | | | | | | | |
|---|---|---|---|---|---|---|---|---|---|---|---|---|---|
| **trt 1** | | | | | | | | | | | | | |
| 0.15<br>(-0.02 - 0.32) | **trt 2** | | | | | | | | | | | | |
| 0.19<br>(-0.04 - 0.42) | 0.04<br>(-0.13 - 0.22) | **trt 3** | | | | | | | | | | | |
| -0.1<br>(-0.44 - 0.24) | -0.25<br>(-0.55 - 0.05) | -0.29<br>(-0.64 - 0.05) | **trt 4** | | | | | | | | | | |
| 0.05<br>(-0.12 - 0.23) | -0.1<br>(-0.2 - 0.01) | -0.14<br>(-0.32 - 0.04) | 0.15<br>(-0.16 - 0.46) | **trt 5** | | | | | | | | | |
| -0.03<br>(-0.23 - 0.17) | -0.18<br>(-0.34 - -0.02) | -0.22<br>(-0.44 - 0) | 0.07<br>(-0.23 - 0.37) | -0.08<br>(-0.24 - 0.08) | **trt 6** | | | | | | | | |
| 0.25<br>(0.05 - 0.44) | 0.1<br>(-0.03 - 0.22) | 0.05<br>(-0.15 - 0.25) | 0.35<br>(0.03 - 0.66) | 0.19<br>(0.07 - 0.32) | 0.27<br>(0.09 - 0.46) | **trt 7** | | | | | | | |
| 0.24<br>(0.06 - 0.43) | 0.09<br>(-0.03 - 0.21) | 0.05<br>(-0.15 - 0.24) | 0.34<br>(0.03 - 0.66) | 0.19<br>(0.06 - 0.32) | 0.27<br>(0.1 - 0.44) | 0<br>(-0.15 - 0.14) | **trt 8** | | | | | | |
| -0.17<br>(-0.73 - 0.4) | -0.32<br>(-0.86 - 0.22) | -0.36<br>(-0.93 - 0.2) | -0.07<br>(-0.68 - 0.55) | -0.22<br>(-0.77 - 0.32) | -0.14<br>(-0.7 - 0.42) | -0.41<br>(-0.96 - 0.13) | -0.41<br>(-0.96 - 0.14) | **trt 9** | | | | | |
| 0.28<br>(0.07 - 0.5) | 0.13<br>(-0.02 - 0.29) | 0.09<br>(-0.13 - 0.31) | 0.38<br>(0.05 - 0.72) | 0.23<br>(0.07 - 0.39) | 0.31<br>(0.11 - 0.52) | 0.04<br>(-0.13 - 0.21) | 0.04<br>(-0.13 - 0.22) | 0.45<br>(-0.1 - 1) | **trt 10** | | | | |
| 0.36<br>(0.14 - 0.58) | 0.21<br>(0.07 - 0.35) | 0.17<br>(-0.06 - 0.4) | 0.46<br>(0.15 - 0.77) | 0.31<br>(0.13 - 0.48) | 0.39<br>(0.19 - 0.59) | 0.12<br>(-0.07 - 0.3) | 0.12<br>(-0.06 - 0.3) | 0.53<br>(-0.03 - 1.09) | 0.08<br>(-0.13 - 0.29) | **trt 11** | | | |
| 0.19<br>(-0.09 - 0.46) | 0.04<br>(-0.19 - 0.27) | -0.01<br>(-0.16 - 0.14) | 0.29<br>(-0.09 - 0.66) | 0.13<br>(-0.1 - 0.37) | 0.22<br>(-0.05 - 0.48) | -0.06<br>(-0.31 - 0.19) | -0.05<br>(-0.3 - 0.19) | 0.35<br>(-0.23 - 0.94) | -0.1<br>(-0.37 - 0.17) | -0.17<br>(-0.45 - 0.1) | **trt 12** | | |
| -0.01<br>(-0.21 - 0.18) | -0.16<br>(-0.29 - -0.04) | -0.21<br>(-0.41 - 0) | 0.09<br>(-0.23 - 0.4) | -0.07<br>(-0.19 - 0.06) | 0.02<br>(-0.16 - 0.19) | -0.26<br>(-0.41 - -0.1) | -0.25<br>(-0.4 - -0.1) | 0.15<br>(-0.4 - 0.71) | -0.3<br>(-0.47 - -0.12) | -0.37<br>(-0.56 - -0.19) | -0.2<br>(-0.45 - 0.05) | **trt 13** | |
| -0.08<br>(-0.3 - 0.14) | -0.23<br>(-0.39 - -0.07) | -0.27<br>(-0.49 - -0.05) | 0.02<br>(-0.3 - 0.34) | -0.13<br>(-0.3 - 0.03) | -0.05<br>(-0.24 - 0.14) | -0.32<br>(-0.51 - -0.14) | -0.32<br>(-0.5 - -0.14) | 0.09<br>(-0.47 - 0.65) | -0.36<br>(-0.56 - -0.17) | -0.44<br>(-0.65 - -0.23) | -0.27<br>(-0.53 - 0) | -0.07<br>(-0.25 - 0.11) | **placebo** |

Figure S4: Forest plots of estimated odds ratios of the 13 labor induction treatment options across five outcomes.

**Outcome2:**
Serious maternal morbidity or death

| | | | | | | | | | | | | | |
|---|---|---|---|---|---|---|---|---|---|---|---|---|---|
| **trt 1** | | | | | | | | | | | | | |
| -0.21 (-0.97 - 0.55) | **trt 2** | | | | | | | | | | | | |
| -0.44 (-1.31 - 0.42) | -0.23 (-0.86 - 0.4) | **trt 3** | | | | | | | | | | | |
| NA | NA | NA | **trt 4** | | | | | | | | | | |
| 0.06 (-0.94 - 1.06) | 0.27 (-0.48 - 1.03) | 0.5 (-0.02 - 1.03) | NA | **trt 5** | | | | | | | | | |
| NA | NA | NA | NA | NA | **trt 6** | | | | | | | | |
| -0.21 (-0.96 - 0.55) | 0.01 (-0.42 - 0.43) | 0.24 (-0.16 - 0.63) | NA | -0.27 (-0.78 - 0.25) | NA | **trt 7** | | | | | | | |
| -0.27 (-1.09 - 0.54) | -0.06 (-0.54 - 0.42) | 0.17 (-0.13 - 0.48) | NA | -0.33 (-0.79 - 0.13) | NA | -0.06 (-0.34 - 0.21) | **trt 8** | | | | | | |
| -0.35 (-1.26 - 0.57) | -0.13 (-0.78 - 0.51) | 0.1 (-0.34 - 0.54) | NA | -0.41 (-0.75 - -0.07) | NA | -0.14 (-0.56 - 0.29) | -0.07 (-0.42 - 0.27) | **trt 9** | | | | | |
| -0.11 (-1.03 - 0.81) | 0.1 (-0.54 - 0.75) | 0.33 (-0.11 - 0.77) | NA | -0.17 (-0.51 - 0.17) | NA | 0.1 (-0.33 - 0.52) | 0.16 (-0.18 - 0.5) | 0.24 (0.24 - 0.24) | **trt 10** | | | | |
| -0.22 (-0.96 - 0.51) | -0.01 (-0.23 - 0.21) | 0.22 (-0.23 - 0.67) | NA | -0.28 (-0.87 - 0.31) | NA | -0.01 (-0.25 - 0.22) | 0.05 (-0.23 - 0.33) | 0.13 (-0.35 - 0.6) | -0.11 (-0.59 - 0.36) | **trt 11** | | | |
| -0.1 (-1.09 - 0.89) | 0.11 (-0.68 - 0.91) | 0.34 (-0.14 - 0.83) | NA | -0.16 (-0.87 - 0.55) | NA | 0.11 (-0.51 - 0.73) | 0.17 (-0.4 - 0.74) | 0.25 (-0.41 - 0.9) | 0.01 (-0.64 - 0.66) | 0.12 (-0.54 - 0.78) | **trt 12** | | |
| -0.15 (-1.32 - 1.02) | 0.06 (-0.87 - 1) | 0.29 (-0.34 - 0.93) | NA | -0.21 (-0.85 - 0.43) | NA | 0.06 (-0.7 - 0.82) | 0.12 (-0.55 - 0.79) | 0.2 (-0.47 - 0.86) | -0.04 (-0.71 - 0.63) | 0.07 (-0.72 - 0.87) | -0.05 (-0.85 - 0.75) | **trt 13** | |
| -0.19 (-1.04 - 0.65) | 0.02 (-0.63 - 0.67) | 0.25 (-0.21 - 0.71) | NA | -0.25 (-0.79 - 0.28) | NA | 0.01 (-0.22 - 0.25) | 0.08 (-0.35 - 0.5) | 0.15 (-0.34 - 0.65) | -0.08 (-0.58 - 0.41) | 0.03 (-0.44 - 0.49) | -0.1 (-0.76 - 0.57) | -0.05 (-0.83 - 0.74) | **placebo** |

Figure S5: Forest plots of estimated odds ratios of the 13 labor induction treatment options across five outcomes.

**Outcome3:**
Serious neonatal morbidity or perinatal death

| | | | | | | | | | | | | | |
|---|---|---|---|---|---|---|---|---|---|---|---|---|---|
| **trt 1** | | | | | | | | | | | | | |
| 0.5<br>(-0.05 - 1.05) | **trt 2** | | | | | | | | | | | | |
| 0.11<br>(-0.43 - 0.65) | -0.4<br>(-0.76 - -0.03) | **trt 3** | | | | | | | | | | | |
| 0.08<br>(-0.45 - 0.62) | -0.42<br>(-0.74 - -0.1) | -0.03<br>(-0.32 - 0.27) | **trt 4** | | | | | | | | | | |
| 0.49<br>(-0.07 - 1.04) | -0.02<br>(-0.38 - 0.35) | 0.38<br>(0 - 0.76) | 0.41<br>(0.05 - 0.76) | **trt 5** | | | | | | | | | |
| 0.03<br>(-0.49 - 0.56) | -0.47<br>(-0.78 - -0.16) | -0.07<br>(-0.36 - 0.22) | -0.05<br>(-0.19 - 0.09) | -0.45<br>(-0.76 - -0.14) | **trt 6** | | | | | | | | |
| -0.04<br>(-0.68 - 0.6) | -0.54<br>(-1.08 - 0) | -0.15<br>(-0.57 - 0.28) | -0.12<br>(-0.57 - 0.33) | -0.53<br>(-1.01 - -0.04) | -0.07<br>(-0.51 - 0.36) | **trt 7** | | | | | | | |
| 0.01<br>(-0.54 - 0.56) | -0.49<br>(-0.84 - -0.14) | -0.1<br>(-0.39 - 0.2) | -0.07<br>(-0.24 - 0.1) | -0.48<br>(-0.89 - -0.06) | -0.02<br>(-0.23 - 0.19) | 0.05<br>(-0.4 - 0.5) | **trt 8** | | | | | | |
| 0.49<br>(-0.13 - 1.1) | -0.01<br>(-0.49 - 0.46) | 0.38<br>(-0.03 - 0.79) | 0.41<br>(0.02 - 0.8) | 0<br>(-0.45 - 0.46) | 0.45<br>(0.07 - 0.84) | 0.53<br>(0.19 - 0.86) | 0.48<br>(0.07 - 0.89) | **trt 9** | | | | | |
| 0.49<br>(-0.04 - 1.03) | -0.01<br>(-0.31 - 0.29) | 0.39<br>(0.07 - 0.7) | 0.41<br>(0.24 - 0.59) | 0.01<br>(-0.32 - 0.34) | 0.46<br>(0.28 - 0.64) | 0.53<br>(0.07 - 1) | 0.48<br>(0.24 - 0.73) | 0.01<br>(-0.38 - 0.39) | **trt 10** | | | | |
| 0.17<br>(-0.41 - 0.74) | -0.33<br>(-0.7 - 0.04) | 0.06<br>(-0.29 - 0.42) | 0.09<br>(-0.23 - 0.41) | -0.32<br>(-0.73 - 0.1) | 0.13<br>(-0.18 - 0.45) | 0.21<br>(-0.19 - 0.61) | 0.16<br>(-0.16 - 0.47) | -0.32<br>(-0.72 - 0.08) | -0.33<br>(-0.66 - 0.01) | **trt 11** | | | |
| 0.45<br>(-0.27 - 1.18) | -0.05<br>(-0.65 - 0.55) | 0.34<br>(-0.14 - 0.83) | 0.37<br>(-0.19 - 0.94) | -0.03<br>(-0.65 - 0.58) | 0.42<br>(-0.14 - 0.98) | 0.49<br>(-0.15 - 1.14) | 0.44<br>(-0.13 - 1.01) | -0.04<br>(-0.67 - 0.6) | -0.04<br>(-0.62 - 0.53) | 0.28<br>(-0.32 - 0.88) | **trt 12** | | |
| -0.1<br>(-0.63 - 0.42) | -0.61<br>(-0.95 - -0.26) | -0.21<br>(-0.54 - 0.12) | -0.18<br>(-0.55 - 0.19) | -0.59<br>(-0.99 - -0.19) | -0.14<br>(-0.49 - 0.22) | -0.06<br>(-0.55 - 0.43) | -0.11<br>(-0.51 - 0.29) | -0.59<br>(-1.06 - -0.13) | -0.6<br>(-0.96 - -0.24) | -0.27<br>(-0.68 - 0.14) | -0.55<br>(-1.14 - 0.03) | **trt 13** | |
| 0.33<br>(-0.15 - 0.81) | -0.17<br>(-0.47 - 0.13) | 0.22<br>(-0.06 - 0.51) | 0.25<br>(-0.02 - 0.52) | -0.15<br>(-0.38 - 0.07) | 0.3<br>(0.07 - 0.53) | 0.37<br>(-0.05 - 0.79) | 0.32<br>(0 - 0.64) | -0.16<br>(-0.55 - 0.24) | -0.16<br>(-0.43 - 0.11) | 0.16<br>(-0.18 - 0.5) | -0.12<br>(-0.68 - 0.44) | 0.43<br>(0.13 - 0.74) | **placebo** |

Figure S6: Forest plots of estimated odds ratios of the 13 labor induction treatment options across five outcomes.

**Outcome4:**
Hyperstimulation (with fetal heart rate changes)

| | | | | | | | | | | | | | |
|---|---|---|---|---|---|---|---|---|---|---|---|---|---|
| **trt 1** | | | | | | | | | | | | | |
| -0.23 (-0.6 - 0.14) | **trt 2** | | | | | | | | | | | | |
| -0.25 (-0.64 - 0.14) | -0.02 (-0.39 - 0.35) | **trt 3** | | | | | | | | | | | |
| NA | NA | NA | **trt 4** | | | | | | | | | | |
| -0.07 (-0.48 - 0.34) | 0.16 (-0.18 - 0.5) | 0.18 (-0.16 - 0.52) | NA | **trt 5** | | | | | | | | | |
| -0.19 (-0.97 - 0.59) | 0.04 (-0.7 - 0.77) | 0.06 (-0.71 - 0.82) | NA | -0.12 (-0.88 - 0.64) | **trt 6** | | | | | | | | |
| -0.5 (-0.91 - -0.1) | -0.28 (-0.61 - 0.06) | -0.26 (-0.62 - 0.11) | NA | -0.43 (-0.73 - -0.14) | -0.31 (-1.06 - 0.43) | **trt 7** | | | | | | | |
| -0.71 (-1.07 - -0.35) | -0.48 (-0.8 - -0.17) | -0.46 (-0.76 - -0.16) | NA | -0.64 (-0.95 - -0.33) | -0.52 (-1.28 - 0.24) | -0.2 (-0.51 - 0.1) | **trt 8** | | | | | | |
| 0.36 (-0.13 - 0.86) | 0.59 (0.22 - 0.96) | 0.61 (0.13 - 1.09) | NA | 0.43 (-0.03 - 0.89) | 0.55 (-0.17 - 1.27) | 0.87 (0.44 - 1.29) | 1.07 (0.63 - 1.51) | **trt 9** | | | | | |
| -0.47 (-0.95 - 0.01) | -0.24 (-0.65 - 0.17) | -0.22 (-0.66 - 0.22) | NA | -0.4 (-0.78 - -0.02) | -0.28 (-1.08 - 0.52) | 0.03 (-0.33 - 0.39) | 0.24 (-0.16 - 0.63) | -0.83 (-1.34 - -0.33) | **trt 10** | | | | |
| -0.33 (-0.81 - 0.15) | -0.1 (-0.5 - 0.3) | -0.08 (-0.5 - 0.34) | NA | -0.26 (-0.68 - 0.17) | -0.14 (-0.92 - 0.65) | 0.18 (-0.21 - 0.57) | 0.38 (-0.03 - 0.79) | -0.69 (-1.16 - -0.21) | 0.15 (-0.33 - 0.62) | **trt 11** | | | |
| -0.76 (-2.08 - 0.56) | -0.53 (-1.85 - 0.79) | -0.51 (-1.77 - 0.75) | NA | -0.69 (-2 - 0.62) | -0.57 (-2.05 - 0.91) | -0.26 (-1.57 - 1.06) | -0.05 (-1.35 - 1.25) | -1.12 (-2.47 - 0.23) | -0.29 (-1.63 - 1.05) | -0.43 (-1.76 - 0.9) | **trt 12** | | |
| 0.07 (-0.34 - 0.47) | 0.29 (-0.04 - 0.63) | 0.31 (-0.05 - 0.68) | NA | 0.13 (-0.13 - 0.4) | 0.26 (-0.47 - 0.98) | 0.57 (0.25 - 0.89) | 0.77 (0.47 - 1.08) | -0.3 (-0.74 - 0.15) | 0.54 (0.13 - 0.95) | 0.39 (-0.03 - 0.82) | 0.83 (-0.49 - 2.14) | **trt 13** | |
| 0.29 (-0.17 - 0.76) | 0.52 (0.12 - 0.92) | 0.54 (0.1 - 0.98) | NA | 0.36 (0 - 0.73) | 0.48 (-0.29 - 1.26) | 0.8 (0.42 - 1.17) | 1 (0.62 - 1.39) | -0.07 (-0.57 - 0.43) | 0.76 (0.35 - 1.18) | 0.62 (0.15 - 1.08) | 1.05 (-0.29 - 2.39) | 0.23 (-0.17 - 0.62) | **placebo** |

Figure S7: Forest plots of estimated odds ratios of the 13 labor induction treatment options across five outcomes.

**Outcome5:**
Vaginal delivery not within 24 hours

| | | | | | | | | | | | | | |
|---|---|---|---|---|---|---|---|---|---|---|---|---|---|
| **trt 1** | | | | | | | | | | | | | |
| 0.23<br>(0 - 0.46) | **trt 2** | | | | | | | | | | | | |
| 0.06<br>(-0.2 - 0.32) | -0.17<br>(-0.43 - 0.09) | **trt 3** | | | | | | | | | | | |
| NA | NA | NA | **trt 4** | | | | | | | | | | |
| -0.25<br>(-0.48 - -0.03) | -0.48<br>(-0.71 - -0.26) | -0.31<br>(-0.56 - -0.06) | NA | **trt 5** | | | | | | | | | |
| -0.25<br>(-0.99 - 0.49) | -0.48<br>(-1.22 - 0.26) | -0.31<br>(-1.06 - 0.44) | NA | 0<br>(-0.7 - 0.71) | **trt 6** | | | | | | | | |
| 0.36<br>(0.09 - 0.63) | 0.13<br>(-0.14 - 0.4) | 0.3<br>(0.03 - 0.58) | NA | 0.62<br>(0.38 - 0.85) | 0.61<br>(-0.13 - 1.36) | **trt 7** | | | | | | | |
| 0.5<br>(0.28 - 0.71) | 0.27<br>(0.03 - 0.51) | 0.44<br>(0.19 - 0.69) | NA | 0.75<br>(0.55 - 0.95) | 0.75<br>(0.01 - 1.48) | 0.13<br>(-0.1 - 0.36)" | **trt 8** | | | | | | |
| -0.46<br>(-1.11 - 0.19) | -0.69<br>(-1.32 - -0.06) | -0.52<br>(-1.18 - 0.14) | NA | -0.21<br>(-0.85 - 0.43) | -0.21<br>(-1.17 - 0.74) | -0.82<br>(-1.45 - -0.2) | -0.96<br>(-1.6 - -0.32) | **trt 9** | | | | | |
| 0<br>(-0.31 - 0.3) | -0.23<br>(-0.55 - 0.08) | -0.06<br>(-0.42 - 0.3) | NA | 0.25<br>(-0.06 - 0.56) | 0.25<br>(-0.52 - 1.01) | -0.36<br>(-0.7 - -0.03) | -0.5<br>(-0.82 - -0.18) | 0.46<br>(-0.22 - 1.14) | **trt 10** | | | | |
| 0.42<br>(-0.13 - 0.96) | 0.19<br>(-0.29 - 0.66) | 0.36<br>(-0.17 - 0.89) | NA | 0.67<br>(0.11 - 1.23) | 0.67<br>(-0.23 - 1.56) | 0.05<br>(-0.54 - 0.65) | -0.08<br>(-0.64 - 0.48) | 0.88<br>(0.07 - 1.69) | 0.42<br>(-0.19 - 1.03) | **trt 11** | | | |
| 0.51<br>(-0.09 - 1.12) | 0.28<br>(-0.32 - 0.89) | 0.46<br>(-0.09 - 1) | NA | 0.77<br>(0.17 - 1.37) | 0.76<br>(-0.16 - 1.69) | 0.15<br>(-0.46 - 0.76) | 0.02<br>(-0.58 - 0.62) | 0.98<br>(0.12 - 1.83) | 0.52<br>(-0.13 - 1.17) | 0.1<br>(-0.66 - 0.86) | **trt 12** | | |
| -1.02<br>(-2.34 - 0.3) | -1.25<br>(-2.53 - 0.03) | -1.08<br>(-2.41 - 0.26) | NA | -0.76<br>(-2.08 - 0.55) | -0.77<br>(-2.26 - 0.72) | -1.38<br>(-2.7 - -0.06) | -1.51<br>(-2.87 - -0.16) | -0.56<br>(-1.99 - 0.88) | -1.02<br>(-2.31 - 0.28) | -1.43<br>(-2.82 - -0.04) | -1.53<br>(-2.97 - -0.09) | **trt 13** | |
| -2.31<br>(-2.71 - -1.92) | -2.54<br>(-2.95 - -2.14) | -2.37<br>(-2.8 - -1.95) | NA | -2.06<br>(-2.43 - -1.7) | -2.06<br>(-2.83 - -1.3) | -2.68<br>(-3.09 - -2.27) | -2.81<br>(-3.21 - -2.42) | -1.85<br>(-2.58 - -1.13) | -2.31<br>(-2.7 - -1.93) | -2.73<br>(-3.38 - -2.08) | -2.83<br>(-3.52 - -2.14) | -1.3<br>(-2.64 - 0.05) | **placebo** |

Figure S8: Ranking of the 8 treatment options for labor induction regarding a utility function which give 30% of weight to cesarean section, 25% to serious maternal morbidity or death, 25% to serious neonatal morbidity or perinatal death, 10% to hypersitimulation and 10% to vagina delivery not within 24 hours. In each plot, the $j$-th bar represents the probabilities of each treatment ranking in the $j$-th place.

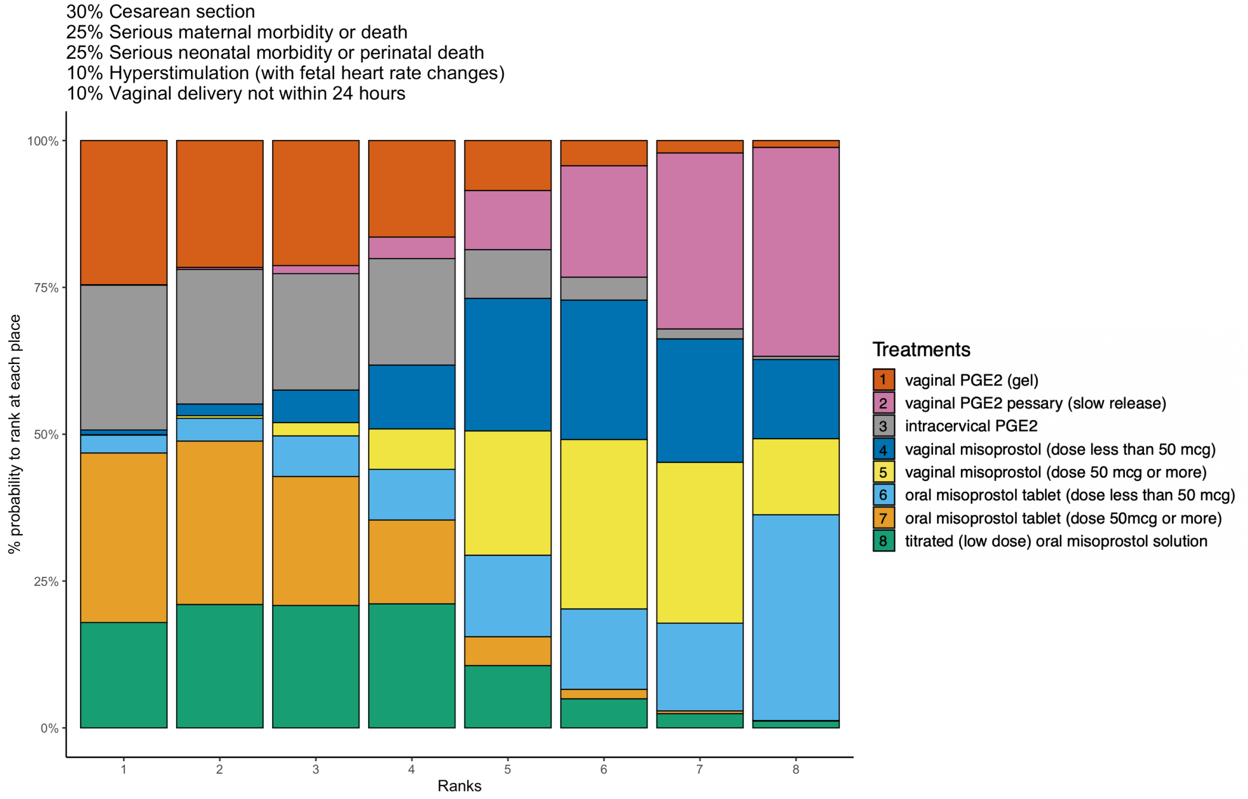

Figure S9: A weight star plot to compare 8 treatment options for labor induction across five outcomes. Outcomes 4 and 5 are given half the weight of outcomes 1, 2, and 3.

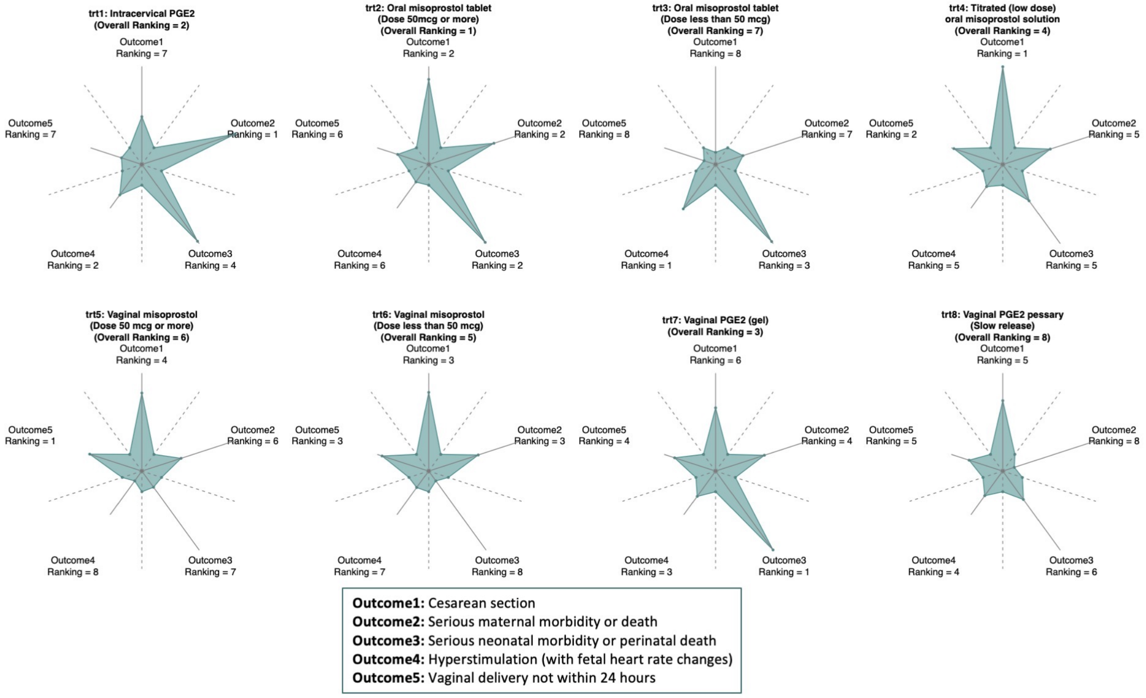

Figure S10: Forest plots of estimated odds ratios of the 13 labor induction treatment options across four outcomes from sensitivity analysis. Due to limited sample size in the network for the outcome maternal morbidity or perinatal death, the outcome was not studied.

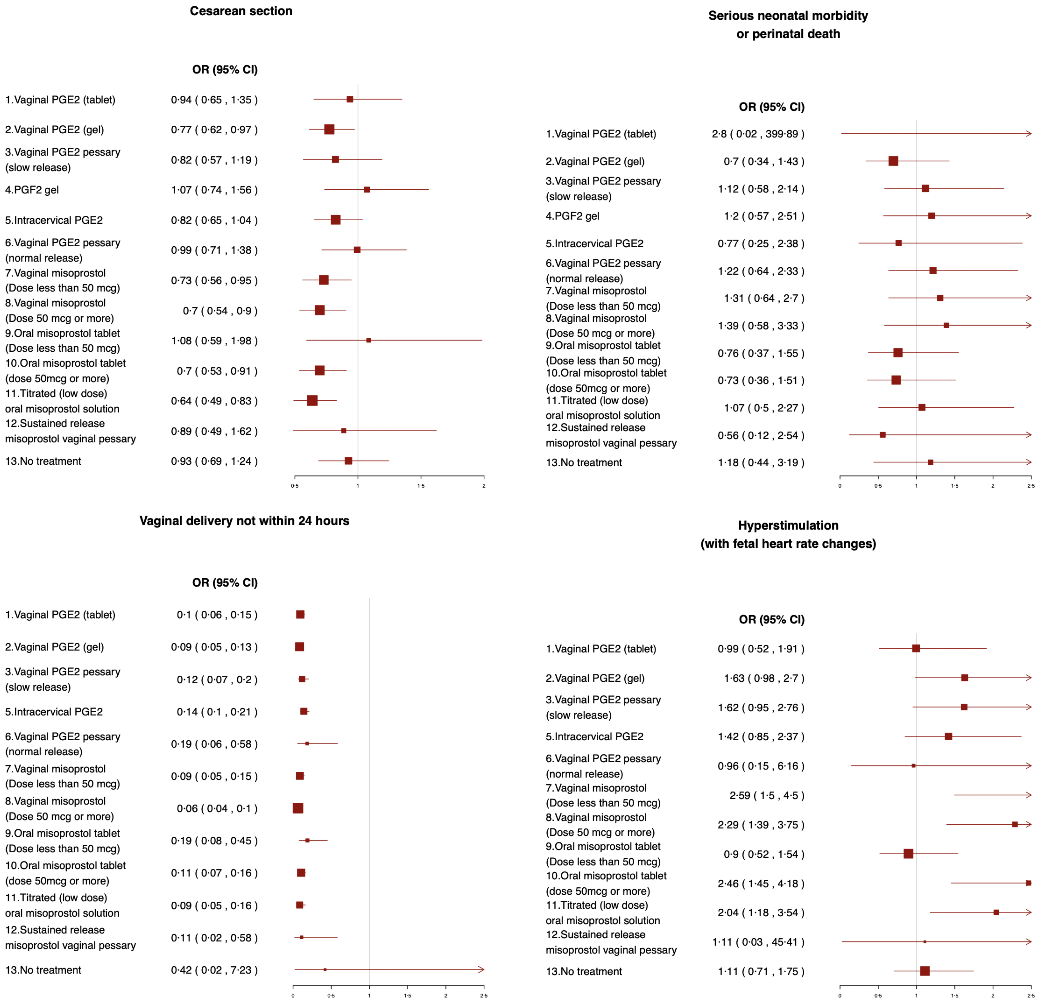

Table S1: WSUCRA of the eight treatments corresponding to the three utility functions. $Y_1$ stands for cesarean section and $Y_2$ stands for maternal morbidity.

| Treatment | $10\%Y_1$ + $90\%Y_2$ | $50\%Y_1$+ $50\%Y_2$ | $90\%Y_1$ + $10\%Y_2$ |
|---|---|---|---|
| vaginal PGE2 (gel) | 0.51 | 0.42 | 0.34 |
| vaginal PGE2 pessary (slow release) | 0.16 | 0.21 | 0.42 |
| intracervical PGE2 | 0.86 | 0.67 | 0.17 |
| vaginal misoprostol (dose less than 50 mcg) | 0.55 | 0.59 | 0.66 |
| vaginal misoprostol (dose 50 mcg or more) | 0.42 | 0.47 | 0.62 |
| oral misoprostol tablet (dose less than 50 mcg) | 0.23 | 0.08 | 0.09 |
| oral misoprostol tablet (dose 50mcg or more) | 0.71 | 0.78 | 0.78 |
| titrated (low dose) oral misoprostol solution | 0.56 | 0.77 | 0.92 |
